# Protocol and Statistical Analysis Plan for the Randomized Trial of Sedative Choice for Intubation (RSI)

**DOI:** 10.1101/2025.01.18.25320768

**Authors:** Stephanie C. DeMasi, Brant Imhoff, Ariel A. Lewis, Kevin P. Seitz, Brian E. Driver, Kevin W. Gibbs, Adit A Ginde, Stacy A. Trent, Derek W. Russell, Amelia L. Muhs, Matthew E Prekker, John P. Gaillard, Daniel Resnick-Ault, L. Jane Stewart, Micah R. Whitson, Graham W. W. Van Schaik, Aaron E. Robinson, Jessica A. Palakshappa, Neil R. Aggarwal, Jason C. Brainard, David J. Douin, Carolynn Lyle, Sheetal Gandotra, Aaron J. Lacy, Kristen C. Sherlin, Greta K. Carlson, J. Maycee Cain, Brianne Redman, Carrie Higgins, Cori Withers, Logan L. Beach, Barbara Gould, Jasmine McIntosh, Bradley D. Lloyd, Tiffany L. Israel, Li Wang, Todd W. Rice, Wesley H. Self, Jin H. Han, Jonathan D. Casey, Matthew W. Semler, the RSI investigators and the Pragmatic Critical Care Research Group

## Abstract

**Background:** Emergency tracheal intubation is a common and high-risk procedure. Ketamine and etomidate are sedative medicines commonly used to induce anesthesia for emergency tracheal intubation, but whether the induction medication used affects patient outcomes is uncertain.

**Research Question:** Does the use of ketamine for induction of anesthesia decrease the incidence of death among adults undergoing emergency tracheal intubation, compared to the use of etomidate?

**Study Design and Methods:** The Randomized trial of Sedative choice for Intubation (RSI) is a pragmatic, multicenter, unblinded, parallel-group, randomized trial being conducted in 14 sites (6 emergency departments and 8 intensive care units) in the United States. The trial compares ketamine vs etomidate for induction of anesthesia among 2,364 critically ill adults undergoing emergency tracheal intubation. The primary outcome is all-cause, 28-day in-hospital mortality. The secondary outcome is the incidence of cardiovascular collapse during intubation, a composite of hypotension, receipt of vasopressors, and cardiac arrest. Enrollment began on April 6, 2022, and is expected to conclude in 2025.

**Interpretation:** The RSI trial will provide important data on the effects of ketamine vs etomidate on death and other outcomes for critically ill adults undergoing emergency tracheal intubation. Specifying the protocol and statistical analysis plan before the conclusion of enrollment increases the rigor, reproducibility, and interpretability of the trial.

**Trial Registry:** ClinicalTrials.gov; No.: NCT05277896; URL: www.clinicaltrials.gov

**Take-Home Points:** *Study Question:* Does use of ketamine for induction of anesthesia during emergency tracheal intubation decrease the incidence of death, compared with use of etomidate?

*Results:* This manuscript describes the protocol and statistical analysis plan for the Randomized trial of Sedative choice for Intubation (RSI) comparing ketamine vs etomidate for induction of anesthesia for emergency tracheal intubation.

*Interpretation:* Prespecifying the full statistical analysis plan before completion of enrollment increases rigor, reproducibility, and transparency of the trial results.

## INTRODUCTION

Millions of critically ill adults undergo emergency tracheal intubation every year worldwide,^1^ approximately 30% of whom die before hospital discharge.^2,3^ Nearly all patients undergoing emergency tracheal intubation in an emergency department (ED) or intensive care unit (ICU) receive a medication to induce anesthesia for the procedure.^4,5^ Whether the medication used to induce anesthesia affects patient outcomes is uncertain.

Etomidate, an imidazole-derived, sedative-hypnotic agent that produces anesthesia by acting on gamma-aminobutyric acid (GABA) receptors, is the medication most often used to induce anesthesia during emergency tracheal intubation in some EDs and ICUs.^5–9^ Etomidate has been described as an “ideal” medication for induction of anesthesia in critically ill adults because of its rapid onset, short duration of action, and limited effect on blood pressure.^10^ However, etomidate inhibits 11-β-hydroxylase in the adrenal glands, which decreases cortisol production and causes adrenal insufficiency for 24 to 72 hours.^11–14^ Among critically ill adults, many of whom experience critical illness-related corticosteroid insufficiency from sepsis or other causes,^15–18^ etomidate-induced adrenal insufficiency has been hypothesized to be associated with organ dysfunction and death.^6^

Ketamine, a dissociative agent that produces anesthesia by acting on N-methyl-D-aspartate (NMDA) receptors, is the medication most often used to induce anesthesia during emergency tracheal intubation in some EDs and ICUs.^8,19^ In contrast to etomidate, ketamine increases cortisol production.^20,21^ However, while ketamine generally increases blood pressure by stimulating the release of catecholamines,^22^ it is a direct negative inotrope^23–26^ and has been reported to precipitate rare episodes of severe hypotension^8,27–29^ or even cardiac arrest, particularly among critically ill patients who have depleted their stores of endogenous catecholamines.^28,30^

Previous small-to-moderate sized randomized trials comparing ketamine and etomidate for emergency tracheal intubation have reported conflicting results, with some finding that use of ketamine decreased short-term mortality^28,31^ and others finding no significant differences in outcomes.^32–34^ Currently, whether use of ketamine decreases mortality for critically ill adults undergoing emergency tracheal intubation compared to use of etomidate remains uncertain.

To address this uncertainty, we designed the <u>R</u>andomized trial of <u>S</u>edative Choice for Intubation (RSI), which will compare ketamine vs etomidate for induction of anesthesia among 2,364 adults undergoing emergency tracheal intubation. We hypothesize that use of ketamine will decrease the incidence of death, compared to etomidate.

## STUDY DESIGN AND METHODS

This article was written in accordance with Standard Protocol Items: Recommendations for Interventional Trials (SPIRIT) guidelines (Table 1 and Section 2 of e-Appendix 1).^35^

**Table 1:** Schedule of Enrollment, Interventions, and Assessments in the RSI trial. Information on the collection of outcomes at 3 and 12 months will be reported in a separate statistical analysis plan. Table entries without data mean that data elements were not collected at that time. RSI = Randomized trial of Sedative Choice during Intubation.

|  | **Eligibility Screen** | **Randomization and Allocation** | **Peri-Procedural** |  |  |  | **Final Outcome Assessment** |
| --- | --- | --- | --- | --- | --- | --- | --- |
| Timepoint | Decision to perform tracheal intubation | Before tracheal intubation | Induction | Tracheal intubation | 0-2 min after tracheal intubation | 0-24 hours after tracheal intubation | 28 days |
| Enrollment: |  |  |  |  |  |  |  |
| Eligibility screen | **X** |  |  |  |  |  |  |
| Enrollment |  | **X** |  |  |  |  |  |
| Allocation |  | **X** |  |  |  |  |  |
| Interventions: |  |  |  |  |  |  |  |
| Ketamine |  |  | **X** |  |  |  |  |
| Etomidate |  |  | **X** |  |  |  |  |
| Screening for contraindications | **X** | **X** | **X** |  |  |  |  |
| Assessments: |  |  |  |  |  |  |  |
| Baseline variables | **X** | **X** |  |  |  |  |  |
| Peri-procedural variables |  | **X** | **X** | **X** | **X** |  |  |
| Adverse events |  | **X** | **X** | **X** | **X** | **X** | **X** |
| Clinical outcomes |  |  |  | **X** | **X** | **X** | **X** |

### Engagement of Patients, Families, and Community Members

Detailed information on engagement with survivors of emergency tracheal intubation, family members, and community members in each phase of the design and conduct of the RSI trial are summarized in Section 3 of e-Appendix 1.

### Study Design

RSI is a pragmatic, multicenter, unblinded, parallel-group, randomized trial comparing the use of ketamine versus etomidate for induction of anesthesia in 2,364 critically ill adults undergoing emergency tracheal intubation in 14 sites (6 EDs and 8 ICUs) in the United States. The primary outcome is all-cause, 28-day in-hospital mortality. The trial is being conducted by the Pragmatic Critical Care Research Group (www.pragmaticcriticalcare.org). The trial was registered prior to initiation of enrollment (ClinicalTrials.gov identifier: NCT05277896).

### Ethics and Regulatory Approval

The RSI trial protocol was approved by the institutional review board (IRB) at Vanderbilt University Medical Center (IRB number: 210500) and the US Food and Drug Administration (FDA) (IND 141424).

The study is being conducted with Exception from Informed Consent Requirements for Emergency Research (EFIC) (21 CFR 50.24).^36^ Plans for community consultation and public disclosure were approved by the single IRB at Vanderbilt University Medical Center. The IRB of each participating site provided local context for the community consultation and public disclosure plan and activities.

Whenever feasible, patients or their legally authorized representatives (LAR) are approached by research personnel to provide prospective, written informed consent to participate in the trial. When prospective, written informed consent is infeasible, patients are enrolled under EFIC and the patient, the patient’s LAR, or a family member is notified of enrollment in the trial at the earliest feasible opportunity (additional details in Section 4 of e-Appendix 1).

### Study Population

Critically ill adults undergoing emergency tracheal intubation with sedation in a participating unit are potentially eligible. Complete lists of inclusion and exclusion criteria are provided in Table 2.

**Table 2:** Inclusion and Exclusion Criteria.

|  |
| --- |
| **Inclusion Criteria** |
| Patient is critically ill and undergoing emergency tracheal intubation with sedation in an enrolling unit |
| Planned procedure is orotracheal intubation using a laryngoscope |
| Planned operator is a clinician expected to routinely perform tracheal intubation in the participating unit |
| **Exclusion Criteria** |
| Patient is known to be less than 18 years old |
| Patient is known to be pregnant |
| Patient is known to be a prisoner |
| Patient is known to have an allergy to ketamine or etomidate |
| Patient is presenting to the emergency department with a primary diagnosis of trauma |
| Patient or LAR declines participation during pre-enrollment opt-out conversation or by wearing opt-out bracelet for the RSI trial |
| Clinician feels ketamine is required or contraindicated for the optimal care of the patient |
| Clinician feels etomidate is required or contraindicated for the optimal care of the patient |
| Clinician feels an induction medication other than ketamine or etomidate is required for the optimal care of the patient |
| Immediate need for intubation precludes safe performance of study procedures |

### Randomization and Treatment Allocation

Patients are randomized in a 1:1 ratio to ketamine or etomidate in permuted blocks of variable size, stratified by study site. Study group assignments are generated using a computerized randomization sequence, placed in sequentially numbered opaque envelopes, and distributed to enrolling sites. Before opening the envelope, the clinician performing the tracheal intubation procedure (referred to as the “operator”) determines that the patient meets eligibility criteria. Study group assignment remains concealed to study personnel and clinicians until after the decision has been made to enroll the patient and the envelope is opened. Patients are enrolled and randomized once the operator opens the trial envelope to reveal study group assignment. After randomization, patients, clinicians, and study personnel are not blinded to trial group assignment.

## STUDY INTERVENTIONS

### Ketamine group

For patients in the ketamine group, clinicians are instructed to administer ketamine intravenously to induce anesthesia for emergency tracheal intubation. The dose of ketamine is determined by the clinicians based on the clinical condition of the patient. A nomogram on the study group assignment sheet provides clinicians with doses of ketamine (in milligrams) that correspond to a full dose (2.0 mg/kg), an intermediate dose (1.5 mg/kg), and a reduced dose (1.0 mg/kg) for a range of patient weights (see Section 5 of e-Appendix 1).^37^

### Etomidate Group

For patients in the etomidate group, clinicians are instructed to administer etomidate intravenously to induce anesthesia for emergency tracheal intubation. The dose of etomidate is determined by the clinicians based on the clinical condition of the patient. A nomogram on the study group assignment sheet provides clinicians with doses of etomidate (in milligrams) that correspond to a full dose (0.3 mg/kg), an intermediate dose (0.25 mg/kg), and a reduced dose (0.2 mg/kg) for a range of patient weights (see Section 6 of e-Appendix 1).

### Co-Interventions

The RSI trial determines only the sedative medication administered for induction of anesthesia during the emergency tracheal intubation procedure. Subsequent boluses or infusions of sedative medications are determined by clinicians. Other aspects of emergency tracheal intubation (e.g., pre-intubation fluid and vasopressor management, choice of neuromuscular blocking medication) are determined by treating clinicians according to clinical protocols in the study units. Co-interventions that could potentially modify the effect of ketamine or etomidate on patient outcomes (e.g., administration of corticosteroids after intubation) are prospectively recorded.

### Data Collection

An observer not directly involved with the intubation procedure collects data in real time for key periprocedural outcomes, including oxygen saturation and systolic blood pressure at the time of induction and lowest oxygen saturation, lowest systolic blood pressure, and administration of vasopressors between induction and two minutes after intubation. Observers are either research personnel or trained clinical personnel (e.g., physicians or nurses). The accuracy of this method of data collection has been validated previously^38^ and used in numerous previous trials of emergency tracheal intubation.^3,5,7^

Immediately after the intubation procedure, the operator completes a paper data collection form to record characteristics of the procedure (e.g., Cormack-Lehane grade of glottic view^39^), complications during intubation (e.g., cardiac arrest), and the operator’s prior intubating experience. Study personnel at each site review the electronic health record to collect data on baseline characteristics, management before and after laryngoscopy, and in-hospital clinical outcomes such as systolic blood pressure at 24 hours after induction, receipt of vasopressors at 24 hours after induction, ventilator duration, and mortality. Study personnel collect information on mortality occurring after hospital discharge from the electronic health record, public vital statistics records and other public records, and phone calls with patients participating in the long-term outcome assessments at 3 and 12 months (the results of the long-term outcome assessments will be reported separately).

### Primary Outcome

The primary outcome is all-cause, 28-day, in-hospital mortality, defined as death from any cause occurring between enrollment and 28 days after enrollment with outcome ascertainment ending at hospital discharge.

### Secondary Outcome

The sole secondary outcome is cardiovascular collapse, defined as the occurrence of any of the following between induction and 2 minutes after intubation: (1) systolic blood pressure < 65 mmHg; (2) new or increased vasopressors; (3) cardiac arrest not resulting in death within 1 hour of induction; (4) cardiac arrest resulting in death within 1 hour of induction.

### Procedural, Clinical, and Safety Outcomes

Table 3 reports the exploratory procedural outcomes, exploratory clinical outcomes, and safety outcomes. Definitions of free-day outcomes are included in Supplemental Section 7 of e-Appendix 1.^5,7^

**Table 3:** Study outcomes.

|  |  |
| --- | --- |
| **Primary Outcome** | All-cause, 28-day, in-hospital mortality |
| **Secondary Outcome** | Cardiovascular collapse, a composite of any of the following between induction and 2 minutes after intubation: (1) systolic blood pressure < 65 mmHg; (2) new or increased vasopressors; (3) cardiac arrest not resulting in death within 1 hour of induction; (4) cardiac arrest resulting in death within 1 hour of induction. |
| **Exploratory Procedural Outcomes** | Cormack-Lehane Grade of glottic view |
|  | Successful intubation on the first attempt, defined as placement of an endotracheal tube in the trachea with a single insertion of a laryngoscope blade into the mouth and EITHER a single insertion of an endotracheal tube into the mouth OR a single insertion of a bougie into the mouth followed by a single insertion of an endotracheal tube over the bougie into the mouth. |
|  | Time from induction to successful tracheal intubation |
|  | Lowest oxygen saturation between induction and two minutes after intubation |
|  | Lowest oxygen saturation < 80% between induction to two minutes after intubation |
|  | Highest and lowest systolic blood pressure from induction to two minutes after intubation |
|  | Systolic blood pressure > 180 between induction and two minutes after intubation |
|  | Systolic blood pressure < 65 mmHg between induction and 2 minutes after intubation |
|  | New or increased vasopressor between induction and 2 minutes after intubation |
|  | Cardiac arrest within 2 minutes of intubation not resulting in death within 1 hour of induction |
|  | Cardiac arrest within 2 minutes of intubation resulting in death within 1 hour of induction |
| **Exploratory Clinical Outcomes** | Ventilator-free days to study day 28 |
|  | Vasopressor-free days to study day 28 |
|  | ICU-free days to study day 28 |
| **Safety Outcomes** | Systolic blood pressure at 24 hours after induction |
|  | Receipt of vasopressors at 24 hours after induction |
|  | Cardiac arrest receiving cardiopulmonary resuscitation between induction and hospital discharge |

### Long-term Outcomes

The definitions, collection, and analysis of survival and functional outcomes (e.g., symptoms of post-traumatic stress disorder) at 3 months and 12 months will be prespecified in a separate statistical analysis plan. Because these outcomes will not be available for more than a year after completion of enrollment in the trial, these long-term outcomes have been registered separately (NCT06179485) and will be reported separately from the 28-day outcomes described in this statistical analysis plan.

### Data and Safety Monitoring Board

The composition and responsibilities of the Data and Safety Monitoring Board (DSMB) are described in Section 8 of e-Appendix 1. The DSMB has the authority to recommend that the trial stop at any point, request additional data, request additional interim analyses, or request modifications to the study protocol.

### Trial Stages

The RSI trial was funded in two stages. First, the single-center, feasibility stage of the trial was funded by an award from the National Heart Lung and Blood Institute (K23HL153584). Second, the multicenter stage of trial was funded by a contract from the Patient-Centered Outcomes Research Institute® (BPS-2022C3-30021). The initial trial protocol specified both stages of the trial and the criteria that had to be met to transition from the single-center stage to the multicenter stage. On August 1, 2023, the DSMB (i) reviewed information on funding for the multicenter stage, (ii) reviewed data on enrollment rate, adherence to trial procedures, and completeness and quality of trial data from the single-center stage, and (iii) approved transition to the multicenter stage of the trial. Neither the DSMB nor the investigators reviewed any analysis of trial outcomes as part of the transition between stages.

### Interim Analysis

On January 7, 2025, the DSMB reviewed a single interim analysis, prepared by the unblinded study biostatistician at the anticipated halfway point of the trial after enrolment of 1,182 patients. The pre-specified stopping boundary for efficacy was a P value < 0.001 for the difference between trial groups in the primary outcome using a generalized linear mixed effects model with a random effect for study site and a fixed effect for group assignment (ketamine group vs etomidate group). This conservative Haybittle–Peto boundary will allow the final analysis to be performed using an unchanged level of significance (two-sided P value < 0.05). After reviewing the interim analysis, the DSMB recommended continuing the RSI trial without modification.

### Sample Size Estimation

The final sample size for the RSI trial was calculated at the time of receipt of funding for the multicenter stage of the trial from PCORI® (details of the initial sample size calculation are provided in Section 9 of e-Appendix 1). Based on prior trials in the same settings,^5,7^ we estimated that the incidence of all-cause, 28-day in-hospital mortality (primary outcome) in the etomidate group would be 30%. The patient and clinician partners recommended that the trial should have adequate statistical power to detect an absolute difference between groups in mortality of approximately 5 percentage points. This difference is more conservative than the median of 8 percentage points (interquartile range, 6-10) used as the minimum clinically important difference in the design of prior randomized trials in critical care^40^ and is smaller than the observed difference in mortality between ketamine and etomidate of 6-7 percentage points observed in two previous trials.^28,32^ We used the *bsamsize* R function to calculate that achieving 80% statistical power at a two-sided alpha of 0.05 to detect a difference in mortality of 5.2 percentage points (30.0% in the etomidate group vs 24.8% in the ketamine group) would require enrollment of 2,308 patients. Anticipating that, like previous EFIC trials,^41^ less than 3% of participants would discontinue follow up before ascertainment of the primary outcome, we planned to enroll a total of 2,364 patients (1,182 per group).

### Statistical Analysis Principles

Analyses will be conducted following reproducible research principles using R (R Foundation for Statistical Computing, Vienna, Austria). Categorical variables will be presented as number and percentage. Continuous variables will be presented as mean ± SD or median (interquartile range). A two-sided P-value of < 0.05 will define a statistically significant between-group difference in the primary outcome. With a single primary outcome, no adjustment for multiplicity will be made. For analyses of secondary and exploratory, emphasis will be placed on the magnitude of differences between groups with 95% confidence intervals, rather than statistical significance. Templates for the tables that will be presented in the results manuscript and supplement for baseline, periprocedural, and in-hospital outcome variables are provided in Supplementary Tables 1-3 in e-Appendix 1.

### Main Analysis of the Primary Outcome

The primary analysis will be an intention-to-treat comparison of patients randomized to the ketamine group versus patients randomized to the etomidate group with regard to the primary outcome of all-cause, 28-day in-hospital mortality among all patients in the trial population except those who withdrew from follow up prior to ascertainment of the primary outcome. The primary outcome will be compared between the two trials groups using a generalized linear mixed effects model with a random effect for study site and a fixed effect for group assignment (ketamine group vs etomidate group). The absolute difference in percentages, associated 95% confidence intervals, and a P value for the comparison will be presented. A rationale for the primary outcome and its analysis can be found in Section 10 of e-Appendix 1.

### Additional Analyses of the Primary Outcome

#### Sensitivity Analyses

We will assess the robustness of the findings of the primary analysis in a series of sensitivity analyses using different approaches to defining and analyzing the primary outcome. First, we will repeat the primary analysis among all patients in the trial population, with patients who withdrew from follow up prior to outcome ascertainment treated as (i) all having experienced the primary outcome or (ii) all having not experienced the primary outcome. Second, we will repeat the primary analysis using all-cause, all-location mortality at 28 days (i.e., including available information on deaths that occur after hospital discharge). Third, we will repeat the primary analysis using a Chi-square test rather than a generalized linear mixed effects model. Fourth, we will compare survival to day 28 between trial groups using the Kaplan-Meier method. Fifth, we will perform an adjusted analysis comparing the primary outcome between groups using a generalized linear mixed effects model with a random effect for trial site and fixed effects for trial group assignment and the following pre-specified baseline variables: age; sex; race; ethnicity; area deprivation index (a proxy for socioeconomic status and an indicator of neighborhood disadvantage);^42^ rurality;^43^ number of comorbidities; and pre-enrollment severity of illness as assessed by the Acute Physiology and Chronic Health Evaluation II score.^44^ Continuous variables will be modelled assuming a non-linear relationship to the outcome using restricted cubic splines with between 3 and 5 knots.

#### Analyses of heterogeneity of treatment effect

Heterogeneity of treatment effect is nonrandom variation in the magnitude or direction of the effect of a treatment on an outcome across levels of a baseline covariate. We will apply the three complementary approaches to analysis of heterogeneity of treatment effect described in the Predictive Approaches to Treatment effect Heterogeneity Statement: (1) traditional one-variable-at-a-time subgroup analyses; (2) a risk-modeling approach; and (3) an effect-modeling approach.^45^ Complete details of all three approaches are described in Section 11 of e-Appendix 1.

Our subgroup analyses will examine whether prespecified baseline variables modify the effect of trial group assignment on the primary outcome using a formal test of statistical interaction in a generalized linear mixed effects model with the primary outcome as the dependent variable, a random effect for trial site, and independent variables of trial group, the proposed effect modifier, and the interaction between the effect modifier and trial group for each of the following prespecified baseline variables:

1. Sepsis or septic shock (Yes / No)
2. Vasopressor receipt (Yes / No)
3. Patient location (ED / ICU)
4. Adrenal insufficiency or chronic receipt of corticosteroids (Yes / No)
5. Acute neurologic condition (Yes / No)
6. Active cardiac condition (Yes /No)
7. Baseline risk of the primary outcome (continuous)

### Analysis of the Secondary Outcome

We will perform an unadjusted, intention-to-treat comparison of patients randomized to the ketamine group versus patients randomized to the etomidate group with regard to the secondary outcome of cardiovascular collapse. A Chi-square test will be used to generate between-group differences and the associated 95% confidence intervals.

### Analyses of Additional Outcomes

We will conduct unadjusted, intention-to-treat analyses comparing patients randomized to the ketamine group versus patients randomized to the etomidate group with regard to each pre-specified exploratory procedural outcome, exploratory clinical outcome, and safety outcome. Between-group differences and the associated 95% confidence intervals will be generated using a Chi-square test for categorical outcomes and a Wilcoxon rank-sum test for continuous outcomes.

### Handling of Missing Data

We anticipate that no data on the primary outcome will be missing except for patients who withdrew from participation prior to the collection of the primary outcome. When data are missing for an outcome, we will perform complete-case analysis, excluding cases where the data for the analyzed outcome are missing. In adjusted analyses, missing data for covariates will be imputed using multiple imputations.

## Discussion

This article reports the rationale, design, and analysis plan for the RSI trial, a 2,364-patient randomized trial of ketamine vs etomidate for induction of anesthesia among critically ill adults undergoing emergency tracheal intubation. Several elements of the trial’s design warrant discussion.

The RSI trial was designed to compare ketamine vs etomidate and does not examine other medications capable of inducing anesthesia for emergency tracheal intubation, such as propofol or benzodiazepines. These comparators were selected for several reasons. First, ketamine and etomidate are the medications most often used for emergency tracheal intubation in many EDs^8^ and ICUs^6^, particularly in the US.^3,46–51^ Second, ketamine and etomidate are the only medications consistently used for emergency tracheal intubation in clinical care at the 14 sites participating in the trial.^5,7,19^ Third, although propofol and benzodiazepines are more commonly used by some clinical specialties (e.g., anesthesiologists)^52,53^ and in some clinical settings (e.g., some ICUs outside of the US),^4^ observational studies have consistently reported these medications to be associated with higher rates of hypotension during intubation, particularly for patients with pre-existing hemodynamic instability.^4,54^ We anticipate that approximately 1-in-4 patients in the RSI trial will be receiving vasopressors before enrollment. Thus, we designed the trial to compare two medications with a hemodynamic profile that permits their use for nearly all critically ill adults, including those with hypotension or shock prior to intubation.

In the RSI trial, clinicians determine what dose of the assigned sedative medication each patient receives based on the clinical condition of the patient using a dosing nomogram provided by the trial. The nomogram specifies 2 mg/kg of ketamine and 0.3 mg/kg of etomidate as a “full dose”. These are the doses specified in the US FDA labeling information for the induction of anesthesia.^37,55^ These are also the doses used in two of the largest previous randomized trials comparing ketamine vs etomidate among critically ill adults.^28,32^ For patients who clinicians determine require a lower dose, the nomogram also provides information on an intermediate dose (1.5 mg/kg of ketamine or 0.25 mg/kg of etomidate) and a reduced dose (1.0 mg/kg of ketamine or 0.2 mg/kg of etomidate) across a range of patient weights.

The design of the RSI trial differs from that of previous trials comparing ketamine vs etomidate in several respects. First, the planned sample size of 2,364 patients is almost three times as large as any prior trial and will permit more precise estimates of treatment effect.^28,32,33,56^ Second, unlike prior trials that excluded patients intubated in the ED,^28^ excluded patients intubated in the ICU,^33^ or enrolled only patients with sepsis,^56^ the RSI trial includes a broad range of patients undergoing emergency tracheal intubation in an ED or ICU, which increases generalizability.^56^ The only diagnosis excluded from the RSI trial is patients presenting to the emergency department with a primary diagnosis of trauma. Because some prior data suggested that ketamine might increase intracranial pressure^57^ and results of one prior trial suggested a potential difference in treatment effect between patients with and without trauma,^32^ we determined that the effect of ketamine vs etomidate on outcomes for patients presenting with trauma was best evaluated in a separate trial. Third, unlike prior trials that did not collect procedural outcomes^32^ or long-term patient-centered outcomes,^28,32,33,56^ the RSI trial collects physiological outcomes during the procedure (e.g., cardiovascular collapse during intubation), short-term patient-centered outcomes (e.g., death by day 28), and long-term patient-centered outcomes (e.g., survival and symptoms of post-traumatic stress disorder at 12 months).

### Interpretation

The RSI trial will provide important evidence regarding the effect of ketamine vs etomidate on death and other clinical outcomes among critically ill adults undergoing emergency tracheal intubation in an ED or ICU. To aid in the transparency and interpretation of trial results, this manuscript detailing the protocol and statistical analysis plan for the RSI trial has been finalized prior to the conclusion of patient enrollment.

## Supporting information

Supplementary file to: Protocol and Statistical Analysis Plan for the Randomized Trial of Sedative Choice for Intubation (RSI)

## Data Availability

n/a (this is a statistical analysis plan)

## Abbreviations

DSMB: Data and Safety Monitoring Board
ED: Emergency Department
EFIC: Exception from Informed Consent Requirements for Emergency Research
FDA: Food and Drug Administration
ICU: Intensive Care Unit
IRB: Institutional Review Board
LAR: Legally Authorized Representative
SPIRIT: Standard Protocol Items: Recommendations for Interventional Trials

## References

1. Kempker JA, Abril MK, Chen Y, Kramer MR, Waller LA, Martin GS. The Epidemiology of Respiratory Failure in the United States 2002-2017: A Serial Cross-Sectional Study. Crit Care Explor 2020;2(6):e0128.

2. Russotto V, Myatra SN, Laffey JG, et al. Intubation Practices and Adverse Peri-intubation Events in Critically Ill Patients From 29 Countries. JAMA 2021;325(12):1164–1172.

3. Casey JD, Janz DR, Russell DW, et al. Bag-Mask Ventilation during Tracheal Intubation of Critically Ill Adults. N Engl J Med 2019;380(9):811–821.

4. Russotto V, Myatra SN, Laffey JG, et al. Intubation Practices and Adverse Peri-intubation Events in Critically Ill Patients From 29 Countries. JAMA 2021;325(12):1164–1172.

5. Gibbs KW, Semler MW, Driver BE, et al. Noninvasive Ventilation for Preoxygenation during Emergency Intubation. N Engl J Med 2024;390(23):2165–2177.

6. Wunsch H, Bosch NA, Law AC, et al. Evaluation of Etomidate Use and Association with Mortality Compared with Ketamine Among Critically Ill Patients. Am J Respir Crit Care Med 2024;

7. Prekker ME, Driver BE, Trent SA, et al. Video versus Direct Laryngoscopy for Tracheal Intubation of Critically Ill Adults. N Engl J Med 2023;389(5):418–429.

8. April MD, Arana A, Schauer SG, et al. Ketamine Versus Etomidate and Peri-intubation Hypotension: A National Emergency Airway Registry Study. Acad Emerg Med Off J Soc Acad Emerg Med 2020;27(11):1106–1115.

9. Driver BE, Semler MW, Self WH, et al. Effect of Use of a Bougie vs Endotracheal Tube With Stylet on Successful Intubation on the First Attempt Among Critically Ill Patients Undergoing Tracheal Intubation: A Randomized Clinical Trial. JAMA 2021;326(24):2488–2497.

10. Etomidate or Midazolam for Rapid Sequence Induction in Patients with Suspected Sepsis? Accessed December 29, 2021. [Internet]. Available from: https://www.jwatch.org/em201012170000001/2010/12/17/etomidate-or-midazolam-rapid-sequence-induction

11. Chan CM, Mitchell AL, Shorr AF. Etomidate is associated with mortality and adrenal insufficiency in sepsis: a meta-analysis*. Crit Care Med 2012;40(11):2945–53.

12. Cuthbertson BH, Sprung CL, Annane D, et al. The effects of etomidate on adrenal responsiveness and mortality in patients with septic shock. Intensive Care Med 2009;35(11):1868–1876.

13. Lipiner-Friedman D, Sprung CL, Laterre PF, et al. Adrenal function in sepsis: The retrospective Corticus cohort study: Crit Care Med 2007;35(4):1012–1018.

14. Tsai M-H, Peng Y-S, Chen Y-C, et al. Adrenal insufficiency in patients with cirrhosis, severe sepsis and septic shock. Hepatol Baltim Md 2006;43(4):673–681.

15. Boonen E, Vervenne H, Meersseman P, et al. Reduced cortisol metabolism during critical illness. N Engl J Med 2013;368(16):1477–1488.

16. Cooper MS, Stewart PM. Corticosteroid insufficiency in acutely ill patients. N Engl J Med 2003;348(8):727–734.

17. Venkatesh B, Finfer S, Cohen J, et al. Adjunctive Glucocorticoid Therapy in Patients with Septic Shock. N Engl J Med 2018;378(9):797–808.

18. Annane D, Renault A, Brun-Buisson C, et al. Hydrocortisone plus Fludrocortisone for Adults with Septic Shock. N Engl J Med 2018;378(9):809–818.

19. Upchurch CP, Grijalva CG, Russ S, et al. Comparison of Etomidate and Ketamine for Induction During Rapid Sequence Intubation of Adult Trauma Patients. Ann Emerg Med 2017;69(1):24–33.e2.

20. Khalili-Mahani N, Martini CH, Olofsen E, Dahan A, Niesters M. Effect of subanaesthetic ketamine on plasma and saliva cortisol secretion. Br J Anaesth 2015;115(1):68–75.

21. Hergovich N, Singer E, Agneter E, et al. Comparison of the effects of ketamine and memantine on prolactin and cortisol release in men. a randomized, double-blind, placebo-controlled trial. Neuropsychopharmacol Off Publ Am Coll Neuropsychopharmacol 2001;24(5):590–593.

22. Spotoft H, Korshin JD, Sørensen MB, Skovsted P. The cardiovascular effects of ketamine used for induction of anaesthesia in patients with valvular heart disease. Can Anaesth Soc J 1979;26(6):463–467.

23. Pagel PS, Kampine JP, Schmeling WT, Warltier DC. Ketamine Depresses Myocardial Contractility as Evaluated by the Preload Recruitable Stroke Work Relationship in Chronically Instrumented Dogs with Autonomic Nervous System Blockade. Anesthesiology 1992;76(4):564–572.

24. Waxman K, Shoemaker WC, Lippmann M. Cardiovascular effects of anesthetic induction with ketamine. Anesth Analg 1980;59(5):355–358.

25. Gelissen HPMM, Epema AH, Henning RH, Krijnen HJ, Hennis PJ, Hertog A den. Inotropic Effects of Propofol, Thiopental, Midazolam, Etomidate, and Ketamine on Isolated Human Atrial Muscle. Anesthesiology 1996;84(2):397–403.

26. Sprung J, Schuetz SM, Stewart RW, Moravec CS. Effects of ketamine on the contractility of failing and nonfailing human heart muscles in vitro. Anesthesiology 1998;88(5):1202–1210.

27. Brown CA, Bair AE, Pallin DJ, Walls RM, NEAR III Investigators. Techniques, success, and adverse events of emergency department adult intubations. Ann Emerg Med 2015;65(4):363–370.e1.

28. Matchett G, Gasanova I, Riccio CA, et al. Etomidate versus ketamine for emergency endotracheal intubation: a randomized clinical trial. Intensive Care Med 2021;

29. Mohr NM, Pape SG, Runde D, Kaji AH, Walls RM, Brown CA. Etomidate Use Is Associated With Less Hypotension Than Ketamine for Emergency Department Sepsis Intubations: A NEAR Cohort Study. Acad Emerg Med Off J Soc Acad Emerg Med 2020;27(11):1140–1149.

30. Dewhirst E, Frazier WJ, Leder M, Fraser DD, Tobias JD. Cardiac arrest following ketamine administration for rapid sequence intubation. J Intensive Care Med 2013;28(6):375–379.

31. Koroki T, Kotani Y, Yaguchi T, et al. Ketamine versus etomidate as an induction agent for tracheal intubation in critically ill adults: a Bayesian meta-analysis. Crit Care Lond Engl 2024;28(1):48.

32. Jabre P, Combes X, Lapostolle F, et al. Etomidate versus ketamine for rapid sequence intubation in acutely ill patients: a multicentre randomised controlled trial. Lancet Lond Engl 2009;374(9686):293–300.

33. Knack SKS, Prekker ME, Moore JC, et al. The Effect of Ketamine Versus Etomidate for Rapid Sequence Intubation on Maximum Sequential Organ Failure Assessment Score: A Randomized Clinical Trial. J Emerg Med 2023;65(5):e371–e382.

34. Greer A, Hewitt M, Khazaneh PT, et al. Ketamine Versus Etomidate for Rapid Sequence Intubation: A Systematic Review and Meta-Analysis of Randomized Trials. Crit Care Med 2024;

35. Chan A-W, Tetzlaff JM, Altman DG, et al. SPIRIT 2013 Statement: Defining Standard Protocol Items for Clinical Trials. Ann Intern Med 2013;158(3):200.

36. 50.24 Exception from informed consent requirements for emergency research. Accessed 11/19/2024.

37. FDA Ketamine. Date Accessed: 11/12/2024 [Internet]. Available from: https://www.accessdata.fda.gov/drugsatfda_docs/label/2018/016812s040lbl.pdf

38. Semler MW, Janz DR, Russell DW, et al. A Multicenter, Randomized Trial of Ramped Position vs Sniffing Position During Endotracheal Intubation of Critically Ill Adults. Chest 2017;152(4):712–722.

39. Cormack RS, Lehane JR, Adams AP, Carli F. Laryngoscopy grades and percentage glottic opening. Anaesthesia 2000;55(2):184–184.

40. Yarnell CJ, Abrams D, Baldwin MR, et al. Clinical trials in critical care: can a Bayesian approach enhance clinical and scientific decision making? Lancet Respir Med [Internet] 2020 [cited 2020 Nov 25];Available from: http://www.sciencedirect.com/science/article/pii/S2213260020304719

41. Silbergleit R, Durkalski V, Lowenstein D, et al. Intramuscular versus intravenous therapy for prehospital status epilepticus. N Engl J Med 2012;366(7):591–600.

42. Kind AJH, Buckingham WR. Making Neighborhood-Disadvantage Metrics Accessible — The Neighborhood Atlas. N Engl J Med 2018;378(26):2456–2458.

43. USDA ERS - Rural-Urban Commuting Area Codes [Internet]. [cited 2021 Dec 14];Available from: https://www.ers.usda.gov/data-products/rural-urban-commuting-area-codes/

44. Knaus WA, Draper EA, Wagner DP, Zimmerman JE. APACHE II: a severity of disease classification system. Crit Care Med 1985;13(10):818–829.

45. Kent DM, Klaveren D van, Paulus JK, et al. The Predictive Approaches to Treatment effect Heterogeneity (PATH) Statement: Explanation and Elaboration. Ann Intern Med 2020;172(1):W1–W25.

46. Ferguson I, Buttfield A, Burns B, et al. Fentanyl versus placebo with ketamine and rocuronium for patients undergoing rapid sequence intubation in the emergency department: The FAKT study-A randomized clinical trial. Acad Emerg Med Off J Soc Acad Emerg Med 2022;29(6):719–728.

47. Ferguson I, Alkhouri H, Fogg T, Aneman A. Ketamine use for rapid sequence intubation in Australian and New Zealand emergency departments from 2010 to 2015: A registry study. Emerg Med Australas EMA 2019;31(2):205–210.

48. Sagarin MJ, Barton ED, Chng Y-M, Walls RM, National Emergency Airway Registry Investigators. Airway management by US and Canadian emergency medicine residents: a multicenter analysis of more than 6,000 endotracheal intubation attempts. Ann Emerg Med 2005;46(4):328–336.

49. Sakles JC, Laurin EG, Rantapaa AA, Panacek EA. Airway management in the emergency department: a one-year study of 610 tracheal intubations. Ann Emerg Med 1998;31(3):325–332.

50. April MD, Long B, Brown CA. Etomidate Should be the Default Agent for Rapid Sequence Intubation in the Emergency Department. Ann Emerg Med 2021;78(6):720–721.

51. Russell DW, Casey JD, Gibbs KW, et al. Effect of Fluid Bolus Administration on Cardiovascular Collapse Among Critically Ill Patients Undergoing Tracheal Intubation: A Randomized Clinical Trial. JAMA 2022;328(3):270–279.

52. Wahlen BM, El-Menyar A, Asim M, Al-Thani H. Rapid sequence induction (RSI) in trauma patients: Insights from healthcare providers. World J Emerg Med 2019;10(1):19–26.

53. Sneyd JR, Gambus PL, Rigby-Jones AE. Current status of perioperative hypnotics, role of benzodiazepines, and the case for remimazolam: a narrative review. Br J Anaesth 2021;127(1):41–55.

54. Wan C, Hanson AC, Schulte PJ, Dong Y, Bauer PR. Propofol, Ketamine, and Etomidate as Induction Agents for Intubation and Outcomes in Critically Ill Patients: A Retrospective Cohort Study. Crit Care Explor 2021;3(5):e0435.

55. FDA Etomidate. Date Accessed: 11/12/2024 [Internet]. 2024;Available from: https://www.accessdata.fda.gov/drugsatfda_docs/label/2017/018227s032lbl.pdf

56. Srivilaithon W, Bumrungphanithaworn A, Daorattanachai K, et al. Clinical outcomes after a single induction dose of etomidate versus ketamine for emergency department sepsis intubation: a randomized controlled trial. Sci Rep 2023;13(1):6362.

57. Zeiler FA, Teitelbaum J, West M, Gillman LM. The ketamine effect on ICP in traumatic brain injury. Neurocrit Care 2014;21(1):163–173.

