## Supplementary file to: Protocol and Statistical Analysis Plan for the Randomized Trial of Sedative Choice for Intubation (RSI) for "Protocol and Statistical Analysis Plan for the Randomized Trial of Sedative Choice for Intubation (RSI)"

### 1. List of RSI Investigators

Coordinating Center: *Clinical Coordinating Center (Vanderbilt University Medical Center, Nashville TN)* – Jonathan D. Casey, MD MSc* (Director, Coordinating Center); Matthew W. Semler, MD MSc* (Chair, Steering Committee); Ariel A. Lewis, MPH, BSN, RN*; Jin H. Han, MD, MSc*; Wesley H. Self, MD, MPH*; Todd W. Rice, MD, MSc*; Christopher G. Hughes, MD; Bradley D. Lloyd, RRT-ACCS*; Karen F. Miller, RN, MPA; Katie S. Gray, BS; Jacob A. Wood, BS; Cheryl L. Gatto, PhD; Mary Lynn Dear, PhD; Grace Van Winkle, BA; Sydney Vidrine, MPH; Tiffany L. Israel, MSSW*; Elizabeth M. Frawley, BSN RN; Madison E. White, RN; Margaret A. Hays, RN MSN.  *Data Coordinating Center (Vanderbilt University Medical Center, Nashville TN)* – Brant Imhoff, MS*; Li Wang, MS*; Matthew S. Shotwell, PhD. *Effect-Modeling Team (University of Wisconsin, Madison, WI)* – Alexandra Spicer MS, Matt Churpek, MD PhD MPH.

Patient Representatives: Aida Strom (Minneapolis, MN); Barbara Gould (Denver, CO)*; Eileen Rubin, (Northbrook, IL); Jasmine McIntosh (Birmingham, AL)*; Patrick Luther (Nashville, TN); Sherman Transou (Winston-Salem, NC).

Denver Health Medical Center: Stacy A. Trent, MD MPH*; Carolynn Lyle, PA-C MPH*; L. Jane Stewart, MD JD MPH*; Marc Retana Castenada, BS; Sonia Iturrino Rijos, BA.

Hennepin County Medical Center: Brian E. Driver, MD*; Aaron E. Robinson, MD MPH*; Matthew E. Prekker, MD MPH*; Greta Kreider Carlson, MD*; Julianna Prohofsky, BS; Joanna Kuo, BS; Kowsar Hurreh, BA; Laurynn Giles, BS; Ananya Narayan, BS; Meghana Chimata, BS, BA; Mia Kellman, BS; Victoria Stehr; Simon Vergara Santibanez, BS; Shawna Ratanpal, BS; Danielle Laibly, BA; Dawson Blankenship, BS; Elijah Hess, BS; Iris Guo; Shivangi Pandey, BS; Spencer Burris-Brown; Stella Frangiadakis.

Wake Forest School of Medicine: Kevin W. Gibbs, MD*; Jessica A. Palakshappa, MD MS*; J. Maycee Cain, BS*; John P. Gaillard, MD*; Brianne Redman, MD*; Haileigh Henson, RN; Savanna Burgess, RN; Benjamin Richards, RN; Belinda Williamson, RN; Lindsey Nicoletti, RN; Summer Usher, RN; Lisa Parks, RN; Leigha Landreth, RN; Katelyn Jimison, PharmD; Alaina Shukdinas, PharmD; Christopher Buckley, PharmD; Karl Healy, PharmD MS; Lars Almassalkhi, PharmD MS; Hannah Morley, PharmD; Adam Furr, PharmD; Adam Boles, PharmD; Robert Dustin Pippen, PharmD.

University of Alabama at Birmingham Medical Center and Heersink School of Medicine: Micah R. Whitson, MD*; Logan L. Beach, MD*; Derek W. Russell, MD*; Sheetal Gandotra, MD*; Mary Clay Boone, RN BSN; Robert B. Johnson, RRT; Geri-Anne Warman, RN BSN; Jennifer J. Oswald, RN BSN; Jerrod Isbell, RRT; Anne Merrill Mason, RN BSN; Gina White, RN BSN; Tyler Greathouse, DO; Morgan Locy, MD PhD; Ryan Goetz, MD; Steven Fox, MD; Jonathan Kalehoff, MD; Daniel Kelmenson, MD; Meena Sridhar, MD; Ahmed Salem, MD; Aneesah B. Jaumally, MD; Ishan Lalani, MD MPH; William S. Stigler, MD; Phillip J. O'Reilly, MD; Donna S. Harris, RN BSN; Cara E. Porter, RN ADN; Sonya Hardy, MA; Puneet Aulakh, MD; Joseph B. Barney, MD;  Joseph Chiles III, MD; Bryan Garcia, MD; Aditya Kotecha, MD; Abdulhakim Tlimat, MD; Peter Morris, MD; Kinner Patel, MD; R. Chad Wade, MD; Carla Copeland, MD; Drew Vestri, RN BSN; Kelsey Jones, RN BSN; Regina Oliver, RN BSN CCRN;  Megan Shelton, MSN RN; Reagan Isbell, RN BSN; Lisa Sarratt, RN BSN; Sarah W. Robison, MD; Nicole Walker, MS RRT; Stephanie Powell, RRT; Great Peagler-Mims, MS RRT; Megan Crumpton, MS RRT;  Laquata Boswell, MS RRT; Dianne Freeman, BS RRT; Sukhmani Boparai, MD; Parker Hambright, MD; Ivan A. Martinez Avalos, MD; Gary Carbell, MD; Nima M. Abdi, MBBS; Hafsa Safdar, MD; Timothy Kennell, MD; Lauren Daley, MD; Victoria Smith, MD; William Smith, MD;  Claudia Tejera Quesada, MD; Hongli Liu, MD PhD; William Nicolson, MD; Kimberly Okoyeze, MD; Andres J. Gonzales Coba, MD PhD; John Craver, MD; K. Cory Guice, DO;  David Page, MD.

University of Colorado School of Medicine: Adit A. Ginde, MD MPH*; Neil R. Aggarwal, MD MHSc*; Carrie Higgins, BSN*; Daniel Resnick-Ault, MD*; Jason C. Brainard, MD*; Cori Withers, BS*; David J. Douin, MD MSc*; Kristine Schauer, MBA RN; Amy Sullivan, BA; Andrew Kluemper, PharmD; Emily Helmer, BS; Kyle Pickard, BS; Laura G. Murphy, BS; Danielle A. Refvem, BSN; Peter Sottile, MD.

Vanderbilt University Medical Center: Stephanie C. DeMasi, MD*; Aaron J. Lacy, MD*; Amelia Muhs, MD*; Graham W. W. Van Schaik MD, MBA*; Kevin P. Seitz, MD MSc*; Kristen C. Sherlin, PharmD*; LaKeysha C. Wiggins, Gabriel A. Kemp, Ryan C. Dillon, PharmD, Jakea D. Johnson, MPH; Linda McLaughlin, PharmD BCPS; Shannon K. Pugh, RN, BSN; Edward T. Qian, MD, MSc; Carla M. Sevin, MD; Joanna L. Stollings, PharmD.

*Denotes members of the writing committee who are listed as authors on the manuscript, the remainder of the RSI investigators represent collaborators.

### 2. SPIRIT 2013 Checklist

SPIRIT 2013 Checklist: Recommended items to address in a clinical trial protocol and related documents*

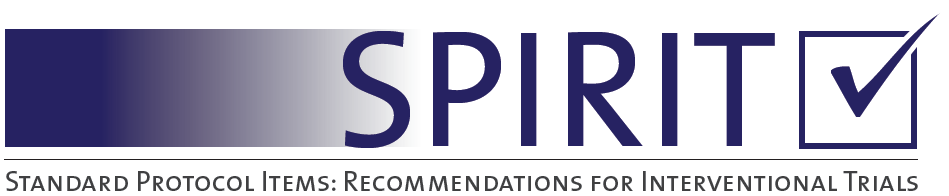

| Section/item | Item  No | | Description | Addressed on page number | |
| --- | --- | --- | --- | --- | --- |
| **Administrative information** | | | |  | |
| Title | 1 | | Descriptive title identifying the study design, population, interventions, and, if applicable, trial acronym | 1 | |
| Trial registration | 2a | | Trial identifier and registry name. If not yet registered, name of intended registry | 4, 7 | |
|  | 2b | | All items from the World Health Organization Trial Registration Data Set | 1-6 | |
| Protocol version | 3 | | Date and version identifier | NA | |
| Funding | 4 | | Sources and types of financial, material, and other support | 2, 12-13 | |
| Roles and responsibilities | 5a | | Names, affiliations, and roles of protocol contributors | 1 | |
|  | 5b | | Name and contact information for the trial sponsor | 2 | |
|  | 5c | | Role of study sponsor and funders, if any, in study design; collection, management, analysis, and interpretation of data; writing of the report; and the decision to submit the report for publication, including whether they will have ultimate authority over any of these activities | 1-3, 12 | |
|  | 5d | | Composition, roles, and responsibilities of the coordinating centre, steering committee, endpoint adjudication committee, data management team, and other individuals or groups overseeing the trial, if applicable (see Item 21a for data monitoring committee) | 1-3  Section 1 of Supplement | |
| Introduction |  | |  |  | |
| Background and rationale | 6a | | Description of research question and justification for undertaking the trial, including summary of relevant studies (published and unpublished) examining benefits and harms for each intervention | 5-6 | |
|  | 6b | | Explanation for choice of comparators | 5-7, 19 | |
| Objectives | 7 | | Specific objectives or hypotheses | 6 | |
| Trial design | 8 | | Description of trial design including type of trial (eg, parallel group, crossover, factorial, single group), allocation ratio, and framework (eg, superiority, equivalence, noninferiority, exploratory) | 7 | |
| Methods: Participants, interventions, and outcomes | | | |  | |
| Study setting | 9 | | Description of study settings (eg, community clinic, academic hospital) and list of countries where data will be collected. Reference to where list of study sites can be obtained | 7, Supplement Section 1 | |
| Eligibility criteria | 10 | | Inclusion and exclusion criteria for participants. If applicable, eligibility criteria for study centres and individuals who will perform the interventions (eg, surgeons, psychotherapists) | Table 2 | |
| Interventions | 11a | | Interventions for each group with sufficient detail to allow replication, including how and when they will be administered | 8-9 | |
|  | 11b | | Criteria for discontinuing or modifying allocated interventions for a given trial participant (eg, drug dose change in response to harms, participant request, or improving/worsening disease) | 8-9 | |
|  | 11c | | Strategies to improve adherence to intervention protocols, and any procedures for monitoring adherence (eg, drug tablet return, laboratory tests) | NA | |
|  | 11d | | Relevant concomitant care and interventions that are permitted or prohibited during the trial | 9 | |
| Outcomes | | 12 | Primary, secondary, and other outcomes, including the specific measurement variable (eg, systolic blood pressure), analysis metric (eg, change from baseline, final value, time to event), method of aggregation (eg, median, proportion), and time point for each outcome. Explanation of the clinical relevance of chosen efficacy and harm outcomes is strongly recommended | | 11-12,  Table 3 |
| Participant timeline | | 13 | Time schedule of enrollment, interventions (including any run-ins and washouts), assessments, and visits for participants. A schematic diagram is highly recommended (see Figure) | | Table 1 |
| Sample size | | 14 | Estimated number of participants needed to achieve study objectives and how it was determined, including clinical and statistical assumptions supporting any sample size calculations | | 13-14, Supplement Section 9 |
| Recruitment | | 15 | Strategies for achieving adequate participant enrollment to reach target sample size | | NA |
| **Methods: Assignment of interventions (for controlled trials)** | | | |  | |
| Allocation: |  | |  |  | |
| Sequence generation | 16a | | Method of generating the allocation sequence (eg, computer-generated random numbers), and list of any factors for stratification. To reduce predictability of a random sequence, details of any planned restriction (eg, blocking) should be provided in a separate document that is unavailable to those who enroll participants or assign interventions | 8 | |
| Allocation concealment mechanism | 16b | | Mechanism of implementing the allocation sequence (eg, central telephone; sequentially numbered, opaque, sealed envelopes), describing any steps to conceal the sequence until interventions are assigned | 8 | |
| Implementation | 16c | | Who will generate the allocation sequence, who will enroll participants, and who will assign participants to interventions | 8 | |
| Blinding (masking) | 17a | | Who will be blinded after assignment to interventions (eg, trial participants, care providers, outcome assessors, data analysts), and how | 8 | |
|  | 17b | | If blinded, circumstances under which unblinding is permissible, and procedure for revealing a participant’s allocated intervention during the trial | NA | |
| **Methods: Data collection, management, and analysis** | | | |  | |
| Data collection methods | 18a | | Plans for assessment and collection of outcome, baseline, and other trial data, including any related processes to promote data quality (eg, duplicate measurements, training of assessors) and a description of study instruments (eg, questionnaires, laboratory tests) along with their reliability and validity, if known. Reference to where data collection forms can be found, if not in the protocol | 10-12 | |
|  | 18b | | Plans to promote participant retention and complete follow-up, including list of any outcome data to be collected for participants who discontinue or deviate from intervention protocols | 14, 18 | |
| Data management | 19 | | Plans for data entry, coding, security, and storage, including any related processes to promote data quality (eg, double data entry; range checks for data values). Reference to where details of data management procedures can be found, if not in the protocol | Supplement Section 14 | |
| Statistical methods | 20a | | Statistical methods for analysing primary and secondary outcomes. Reference to where other details of the statistical analysis plan can be found, if not in the protocol | 14-17, Supplement Section 10, 11 | |
|  | 20b | | Methods for any additional analyses (eg, subgroup and adjusted analyses) | 14-17, Supplement Section 11 | |
|  | 20c | | Definition of analysis population relating to protocol non-adherence (eg, as randomised analysis), and any statistical methods to handle missing data (eg, multiple imputation) | 14-17 | |
| **Methods: Monitoring** | | | |  | |
| Data monitoring | 21a | | Composition of data monitoring committee (DMC); summary of its role and reporting structure; statement of whether it is independent from the sponsor and competing interests; and reference to where further details about its charter can be found, if not in the protocol. Alternatively, an explanation of why a DMC is not needed | Supplement Section 8 | |
|  | 21b | | Description of any interim analyses and stopping guidelines, including who will have access to these interim results and make the final decision to terminate the trial | 12 | |
| Harms | 22 | | Plans for collecting, assessing, reporting, and managing solicited and spontaneously reported adverse events and other unintended effects of trial interventions or trial conduct | Supplement Section 12 | |
| Auditing | 23 | | Frequency and procedures for auditing trial conduct, if any, and whether the process will be independent from investigators and the sponsor | Supplement Section 12 | |
| Ethics and dissemination | | | |  | |
| Research ethics approval | 24 | | Plans for seeking research ethics committee/institutional review board (REC/IRB) approval | 7-8 | |
| Protocol amendments | 25 | | Plans for communicating important protocol modifications (eg, changes to eligibility criteria, outcomes, analyses) to relevant parties (eg, investigators, REC/IRBs, trial participants, trial registries, journals, regulators) | Supplement Section 13 | |
| Consent or assent | 26a | | Who will obtain informed consent or assent from potential trial participants or authorised surrogates, and how (see Item 32) | Supplement Section 4 | |
|  | 26b | | Additional consent provisions for collection and use of participant data and biological specimens in ancillary studies, if applicable | NA | |
| Confidentiality | 27 | | How personal information about potential and enrolled participants will be collected, shared, and maintained in order to protect confidentiality before, during, and after the trial | Supplement Section 14 | |
| Declaration of interests | 28 | | Financial and other competing interests for principal investigators for the overall trial and each study site | 1-3 | |
| Access to data | 29 | | Statement of who will have access to the final trial dataset, and disclosure of contractual agreements that limit such access for investigators | NA | |
| Ancillary and post-trial care | 30 | | Provisions, if any, for ancillary and post-trial care, and for compensation to those who suffer harm from trial participation | Supplement Section 17 | |
| Dissemination policy | 31a | | Plans for investigators and sponsor to communicate trial results to participants, healthcare professionals, the public, and other relevant groups (eg, via publication, reporting in results databases, or other data sharing arrangements), including any publication restrictions | Supplement Section 3 | |
|  | 31b | | Authorship eligibility guidelines and any intended use of professional writers | NA | |
|  | 31c | | Plans, if any, for granting public access to the full protocol, participant-level dataset, and statistical code | NA | |
| Appendices |  | |  |  | |
| Informed consent materials | 32 | | Model consent form and other related documentation given to participants and authorized surrogates | Supplement Section 17 | |
| Biological specimens | 33 | | Plans for collection, laboratory evaluation, and storage of biological specimens for genetic or molecular analysis in the current trial and for future use in ancillary studies, if applicable | NA | |

*It is strongly recommended that this checklist be read in conjunction with the SPIRIT 2013 Explanation & Elaboration for important clarification on the items. Amendments to the protocol should be tracked and dated. The SPIRIT checklist is copyrighted by the SPIRIT Group under the Creative Commons “[Attribution-NonCommercial-NoDerivs 3.0 Unported](http://www.creativecommons.org/licenses/by-nc-nd/3.0/)” license.

### 3. Engagement of Patients, Families, and Community Members

The RSI trial has engaged patients, families, and community members in every phase of the research. Prior to beginning the research, community engagement studios with more than 75 unique patient, family, community member, and clinician partners first identified the choice between ketamine and etomidate as a critical evidence gap and identified mortality as the most relevant and important short-term outcomes and symptoms of post-traumatic stress disorder (PTSD) as the most important non-mortality long-term outcome.

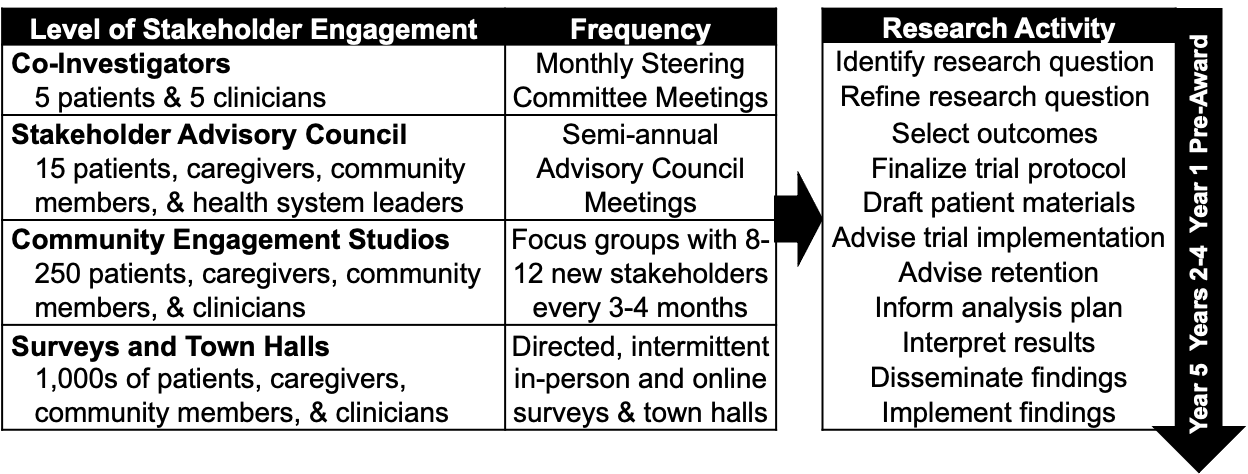
Throughout the conduct of the RSI trial, engagement with patients, families, and community partners occurs at the four levels of engagement shown in the adjacent figure.

Six **Patient Co-investigators**, 5 of whom are survivors of emergency tracheal intubation, serve on the Steering Committee for the RSI trial, contributing to each monthly meeting as equal partners with equal votes. In this role, they have applied their experience building community-research partnerships to the design of pre-trial Community Consultation, their expertise in lay presentation of medical information to optimize the RSI trial’s patient-facing materials and website, and the acceptability to ICU survivors of the approach to informed consent and outcome assessment. At trial completion, the patient co-investigators will help write publications, design dissemination materials, and present the results of the trial to lay and medical audiences.

A **Stakeholder Advisory Council**, composed of 15 patient and community stakeholders knowledgeable about critical illness and representative of the full diversity of their communities, helped design the trial protocol, and meets twice a year with investigators to monitor the impact of the trial on participants, review patient-facing materials, inform participant compensation, and identify opportunities to disseminate results through community organizations and health systems.

In a **Community Engagement Studio**, a unique panel of 8-12 patients, community members, or clinicians (selected for their firsthand knowledge of a particular condition) serve as paid experts during a 2-hour face-to-face session with researchers. During the RSI trial, thrice-yearly Community Engagement Studios focus on: Community Consultation; integration of the trial into clinical care, retention, and long-term outcome assessments; analysis plan development; and interpretation of results, dissemination, and implementation.

Through **Surveys and Town Halls**, we have received input on the RSI trial from hundreds of patients, caregivers, clinicians, and community members. During pre-trial Community Consultation, we conducted in-person surveys with almost 800 patients and family members in ED and ICU waiting rooms to solicit input on the conduct of the RSI trial. ICU survivors, caregivers, and community members in our town halls shared what outcomes they valued, how to make enrollment represent them, and how to communicate with patients about the trial. Through Facebook ads, we solicited input from >1.1 million community members on the trial design. The full details of the Community Consultation and Public Disclosure processes for the RSI trial are publicly available through FDA Docket Number 95S-0158 in the Division of Dockets Management (HFA-305) (<https://www.regulations.gov/document/FDA-1995-S-0036-0230>). The details of the Community Consultation and Public Disclosure processes and the feedback received from patients and community members for the RSI trial will be published separately.

### 4. Ethics and Informed Consent Processes

#### 4.1 Exception from Informed Consent Requirements for Emergency Research (EFIC)

The RSI trial has been approved by the institutional review board at Vanderbilt University Medical Center (IRB number: 210500) and the US Food and Drug Administration (IND 141424). The RSI trial meets all the requirements to enroll patients under 21 CFR 50.24 for emergency research in which prospective informed consent is infeasible.^1^ These requirements include:

1. Human subjects are in a life-threatening situation, available treatments are unproven or unsatisfactory
2. Obtaining informed consent is not feasible
3. Participation in the research holds out the prospect of direct benefit to the subjects
4. The clinical investigation could not practicably be carried out without the waiver
5. The investigator has committed to attempting to contact a legally authorized representative for each subject within that window of time and, if feasible, to asking the legally authorized representative contacted for consent within that window rather than proceeding without consent
6. The IRB has reviewed and approved informed consent procedures and an informed consent document consistent with 50.25; and
7. Additional protections of the rights and welfare of the subjects are provided.

Additional protections of the rights and welfare of the subjects in the RSI trial include:

- Completion of consultation with representatives of the communities in which the clinical investigation will be conducted and from which the subjects will be drawn (community consultation)
- Completion of public disclosure to the communities in which the clinical investigation will be conducted and from which the subjects will be drawn, prior to initiation of the clinical investigation, of plans for the investigation and its risks and expected benefits
- A plan for public disclosure of sufficient information following completion of the clinical investigation to apprise the community and researchers of the study, including the demographic characteristics of the research population, and its results
- Establishment of an independent data monitoring committee to exercise oversight of the clinical investigation; and
- A plan, when feasible, to provide an opportunity for patients, LARs, or family members to object to the subject's participation in the clinical investigation when informed consent is not feasible;

A summary of the pre-trial community consultation and public disclosure activities is publicly available through FDA Docket Number 95S-0158 in the Division of Dockets Management (HFA-305) (https://www.regulations.gov/document/FDA-1995-S-0036-0230).

#### 4.2 Informed Consent Processes in the RSI trial

As required under 21 CFR 50.24,^1^ the RSI trial uses three approaches to informed consent, tailored to the capacity of the patient and the urgency of the intubation procedure.


##### *4.2.1 Written Informed Consent*

When a patient has decisional capacity or a legally authorized representative (LAR) is available and the clinical urgency of the procedure permits sufficient time for trial personnel to complete an informed consent process, written informed consent is obtained from the patient or LAR by trial personnel. The decisional capacity of the patient and the safety and feasibility of providing an opportunity for patients or LARs to complete an informed consent process is determined by treating clinicians.

##### *4.2.2 Opportunity to Decline Participation*

The RSI trial provides two mechanisms for patients to decline participation.  First, medical alert bracelets have been made available prior to and throughout the trial for any community members who wishes to opt out of participation in the trial, should they become critically ill and require tracheal intubation.

Second, when the clinical urgency of the intubation procedure precludes completion of a full informed consent process but permits a brief discussion of the research prior to the procedure, research personnel provide patients, LARs, or family members with an opportunity to express their objection to enrollment in the trial.  The decisional capacity of the patient and the safety and feasibility of providing an opportunity for patients, LARs, or family members to express their objection to research enrollment is determined by treating clinicians.

##### *4.2.3 Exception from Informed Consent*

Because most critically ill patients undergoing emergency tracheal intubation lack decisional capacity due to their critical illness, LARs and family members are frequently unavailable, and tracheal intubation is a time-sensitive procedure with only minutes between the decision to intubate and the completion of the procedure, most patients in the RSI trial are enrolled under EFIC. When a patient is enrolled under EFIC, research personnel notify the patient, LAR, or a family member of the patient’s enrolment in the trial at the earliest feasible opportunity and provide an opportunity to discontinue participation in the trial.

### 5. Group Assignment Sheets with Nomogram (Ketamine)

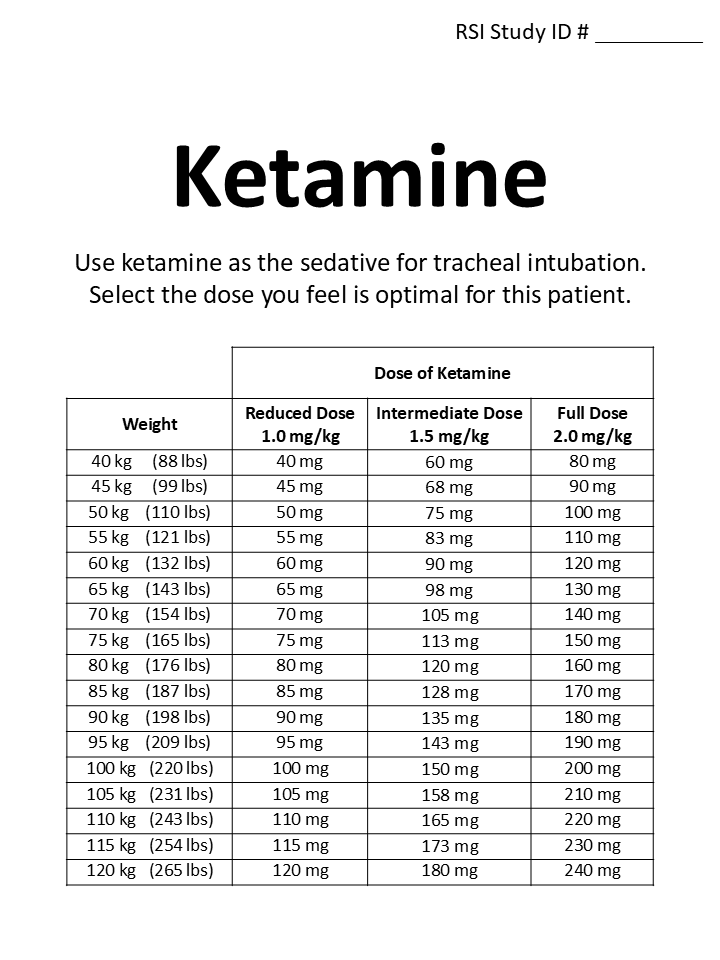

### 6. Group Assignment Sheets with Nomogram (Etomidate)

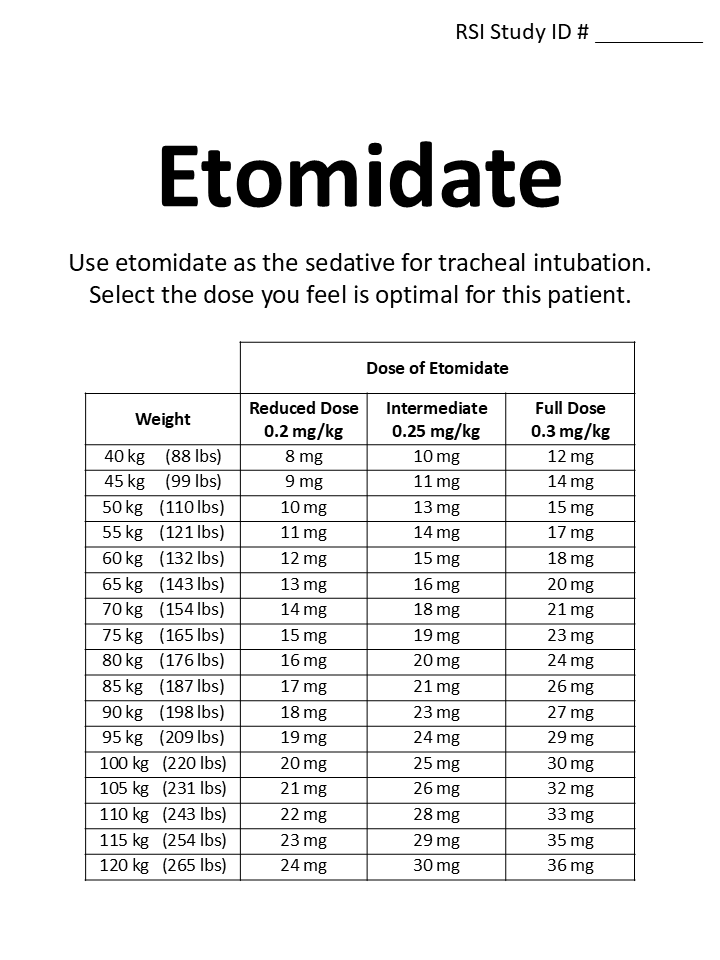

### 7. Definitions of Free-Day Outcomes

*Ventilator-free days to day 28 (VFDs):* VFDs are defined as the number of calendar days, between enrollment and 28 days after enrollment, on which the patient is alive and free of invasive mechanical ventilation. If a patient is liberated from invasive mechanical ventilation, returns to invasive mechanical ventilation and subsequently is liberated from invasive mechanical ventilation again prior to day 28, the number of VFDs will be counted from the end of the last period of invasive mechanical ventilation to day 28. If the patient is receiving invasive mechanical ventilation at day 28 or dies prior to day 28, VFDs are 0. If a patient is discharged while receiving invasive mechanical ventilation, VFDs are 0. Outcome ascertainment ends at 28 days or hospital discharge, whichever occurs first.

*ICU-free days to day 28 (ICU-FDs):* ICU-FDs are defined as the number of calendar days, between enrollment and 28 days after enrollment, on which the patient is alive and not admitted to an intensive care unit after the patient’s final transfer out of the intensive care unit. Patients who are never transferred out of the intensive care unit receive a value of 0. Patients who die before day 28 receive a value of 0. For patients who are transferred out of the ICU, return to an ICU, and are subsequently transferred out of the ICU again prior to day 28, ICU-free days are counted from the date of final transfer out of the ICU. Outcome ascertainment ends at 28 days or hospital discharge, whichever occurs first.

*Vasopressor-free days to study day 28:* Vasopressor-free days are defined as the number of calendar days, between enrollment and 28 days after enrollment, on which the patient is alive and not receiving vasopressors including days before the first receipt of vasopressors and after the last receipt of vasopressors. If a patient is weaned off vasopressors, returns to receiving vasopressors, and is subsequently weaned off vasopressors again, vasopressors free days will not include any days between the first and last receipt of vasopressors. If the patient is receiving vasopressors at day 28 or dies prior to day 28, vasopressor-free days are 0. If a patient is discharged while receiving vasopressors (e.g., transfer to long-term acute care hospital), vasopressors-free days are 0. Outcome ascertainment ends at 28 days or hospital discharge, whichever occurs first.

### 8. Composition and Responsibilities of the Data Safety Monitoring Board

The Data and Safety Monitoring Board (DSMB) consists of members with expertise in bioethics, emergency medicine, pulmonary and critical care medicine, anesthesia, biostatistics, and clinical trials. One patient partner who is a survivor of critical illness requiring emergency tracheal intubation serves as a member of the DSMB. All members of the DSMB are voting members. The DSMB developed a charter, approved the trial protocol, approved the patient notification forms, and approved the trial monitoring plan prior to trial initiation of enrollment. The DSMB has the ability at any point to recommend that the trial end, be modified, or continue unchanged.

The principal role of the DSMB is to assure the safety of patients in the trial. The DSMB monitors data from the trial at semi-annual meetings, reviews and assesses the performance of its operations, and makes recommendations to the Steering Committee and Sponsor with respect to:

- Review of adverse events
- Interim results of the study for evidence of efficacy or adverse events
- Possible early termination of the trial because of new external information, early attainment of study objectives, safety concerns, or inadequate performance
- Possible modifications in the clinical trial protocol
- Performance of individual centers

### 9. Details of Sample Size Calculation

Initial Sample Size Estimate

The initial version of the sample size estimate, included in version 1.0 of the trial protocol submitted on March 9, 2022, is described here.

To estimate the expected incidence of in-hospital mortality in the etomidate group of the RSI trial, we used data from 656 adults undergoing tracheal intubation for non-traumatic critical illness who were recently enrolled from EDs and ICUs in two recent randomized trials.^2,3^ Among these patients, the incidence of 28-day in-hospital mortality was 31%. The minimum clinically important difference (MCID) in mortality used in the design of prior critical care RCTs was an absolute risk reduction of a median of 8 percent (IQR, 6-10).^4^ We powered the RSI trial to detect a more conservative absolute risk reduction of 6 percent. This equated to an incidence of the primary outcome of 25% in the ketamine group and a relative risk of mortality with ketamine compared to etomidate of 0.81 – comparable to the relative risk of 0.827 among adults with non-traumatic critical illness observed in the only prior RCT comparing ketamine vs etomidate.^5^ Achieving 80% statistical power at a two-sided alpha of 0.05 to detect a 6 percent absolute difference between groups in the primary outcome would require enrolling 911 patients per group (1,822 overall). Anticipating that, like prior Exception from Informed Consent Requirements for Emergency Research (EFIC) trials, less than 5% of patients would discontinue participation after enrollment, we planned to enroll a total of 1,900 patients.

Final Sample Size Estimate

The final version of the sample size estimate, included in the trial protocol on October 18, 2023, at the time of funding of the multicenter stage of the RSI trial, is described here.

At the time of funding of the multicenter stage of the RSI trial through a Patient-Centered Outcomes Research Institute® Award (BPS-2022C3-30021), the sample size was re-estimated using updated estimates of mortality and discontinuation of participation by patients. Based on two newly completed trials in the same settings,^2,3^ we updated our estimate for the incidence of all-cause, 28-day in-hospital mortality (primary outcome) in the etomidate group to 30%. The patient and clinician partners who participated in the development of the PCORI® application also recommended that, for two common and inexpensive interventions, the trial should have adequate statistical power to detect an absolute difference between groups in mortality of approximately 5 percentage points. We used the *bsamsize* R function to calculate that achieving 80% statistical power at a two-sided alpha of 0.05 to detect a difference in mortality of 5.2 percentage points (30.0% in the etomidate group vs 24.8% in the ketamine group) would require enrollment of 2,308 patients. Anticipating that, like prior EFIC trials, less than 3% of participants would discontinue follow up before ascertainment of the primary outcome, we planned to enroll a total of 2,364 patients (1,182 per group).

### 10. Rationale for the Choice and Analysis of the Primary Outcome

Patients, caregivers, and clinicians on our Steering Committee and in our town halls, Community Engagement Studios, and surveys consistently identified death as a relevant and important outcome of critical illness. Similarly, survival was a universally-recommended outcome in recent international surveys of patients,^6–9^ clinicians,^10^ and professional societies.^11,12^ The 1-month (28 day) timeframe was chosen because any effects of ketamine vs etomidate on the risk of death likely occur over days to weeks, and the risk of death from comorbidities or new health conditions unrelated to the initial critical illness increases over time.

The choice to analyze mortality as a binary variable rather than a time-to-event analysis was based on input from patient stakeholders who reported that, for patients on a breathing machine, death after a prolonged period on a breathing machine did not represent a better outcome than dying sooner.^13^ This position is consistent with those expressed by patients and caregivers in prior studies.^14–16^

### 11. Effect Modification

We will apply the three complementary approaches to analysis of heterogeneity of treatment effect described in the Predictive Approaches to Treatment effect Heterogeneity (PATH) Statement: (1) traditional one-variable-at-a-time subgroup analyses; (2) a risk-modeling approach; and (3) an effect-modeling approach.

Subgroup Analyses. We will examine whether prespecified baseline variables modify the effect of trial group assignment on the primary outcome using a formal test of statistical interaction in a generalized linear mixed effects model with the primary outcome as the dependent variable, a random effect for trial site, and independent variables of trial group, the proposed effect modifier, and the interaction between the effect modifier and trial group. For categorical variables, we will present the absolute difference and 95% CIs within each prespecified subgroup. Continuous variables will not be dichotomized for analysis of effect modification but may be dichotomized for data presentation. In accordance with the Instrument for assessing the Credibility of effect Modification Analyses (ICEMAN) recommendations,^17^ we have prespecified the following baseline variables as potential effect modifiers and hypothesized the direction of effect modification for each:

1. Sepsis or septic shock (Yes vs No): We hypothesize that the presence of sepsis or septic shock at enrollment, assessed using the Third International Consensus Definitions for Sepsis and Septic Shock (Sepsis-3) definition,^18^ will modify the effect of trial group assignment on the primary outcome, with a greater decrease in 28-day in-hospital mortality in the ketamine group compared to the etomidate group among patients with sepsis or septic shock, compared to patients without sepsis or septic shock. This hypothesis is supported by inconsistent evidence from prior clinical trials and observational studies suggesting that etomidate-induced decreases in cortisol production may have a larger detrimental effect on outcomes for patients with sepsis or septic shock.^19–26^
2. Vasopressor receipt (Yes vs No): We hypothesize that receipt of vasopressors in the hour prior to enrollment will modify the effect of trial group assignment on the primary outcome, with a greater decrease in 28-day in-hospital mortality in the ketamine group compared to the etomidate group among patients receiving vasopressors, compared to patients not receiving vasopressors. This hypothesis is supported by secondary outcomes of prior randomized trials and observational studies that suggest etomidate may worsen shock through causing adrenal insufficiency.^26^
3. Patient location (ED vs ICU): We hypothesize that patient location at enrollment will not modify the effect of trial group assignment on the primary outcome.
4. Adrenal insufficiency or chronic receipt of corticosteroids (Yes vs No): We hypothesize that a pre-existing diagnosis of adrenal insufficiency or chronic receipt of corticosteroids prior to enrollment will modify the effect of trial group assignment on the primary outcome, with a greater decrease in 28-day in-hospital mortality in the ketamine group compared the etomidate group among patients with adrenal insufficiency or chronic receipt of corticosteroids, compared to patients without adrenal insufficiency or chronic receipt of corticosteroids. This hypothesis is supported by mechanistic information on the effect of etomidate on synthesis of cortisol by the adrenal glands.^27^ In prior observational and interventional studies, the administration of corticosteroids during the acute illness has not appeared to affect the association between receipt of etomidate and death.^19,28^
5. Acute neurologic condition (Yes vs. No). We hypothesize that the presence at the time of enrollment of an acute neurologic condition associated with an increased risk for elevated intracranial pressure, defined as intracranial bleeding, meningitis or encephalitis, or stroke, will modify the effect of trial group assignment on the primary outcome, with a lesser decrease in 28-day in-hospital mortality in the ketamine group compared to the etomidate group among patients with an acute neurologic condition, compared to patients without an acute neurologic condition. This hypothesis is supported by inconsistent evidence that ketamine may increase intracranial pressure in some patients.^29^
6. Active cardiac condition (Yes vs. No). We hypothesize that the presence at the time of enrollment of an active cardiac condition, defined as cardiac arrest, cardiogenic shock, congestive heart failure, cardiogenic pulmonary edema, pulmonary hypertension, or myocardial infarction, will modify the effect of trial group assignment on the primary outcome, with a lesser decrease in 28-day in-hospital mortality in the ketamine group compared to the etomidate group among patients with an active cardiac condition, compared to patients without an active cardiac condition. This hypothesis is supported by inconsistent pre-clinical evidence that ketamine may exert a negative ionotropic effect.^30,31^
7. Baseline risk of the primary outcome (continuous): We hypothesize that increased risk of death (increased severity of illness), will modify the effect of trial group assignment on the primary outcome, with a greater decrease in 28-day in-hospital mortality in the ketamine group compared the etomidate group among patients at higher risk of death. This hypothesis is supported by prior research suggesting that patients in critical care trials at higher risk of an outcome frequently experience a larger absolute treatment effect.^32^

Risk-modeling approach. We will examine whether patients’ baseline risk of the primary outcome modifies the effect of trial group assignment on the primary outcome using a formal test of statistical interaction in a generalized linear mixed effects model with the primary outcome as the dependent variable, a random effect for trial site, and independent variables of trial group, patients’ baseline risk of the primary outcome, and the interaction between baseline risk of the primary outcome and trial group. Patients’ baseline risk of the primary outcome will be defined using two approaches. First, a previously derived and validated multivariable model that uses patient characteristics at the time of intubation to predict the probability of 28-day in-hospital mortality for critically ill adults will be used to generate a predicted probability of the primary outcome (ranging from 0.0 to 1.0) for each patient in the trial. Second, each patient’s value at enrolment for the APACHE II score will be used as a measure of the baseline risk.^33^

Effect-modeling approach. The goal of our effect-modeling analyses is to estimate the effect of ketamine vs etomidate on 28-day in-hospital mortality for an individual patient based on his or her baseline characteristics considered simultaneously (“individualized treatment effect”).^34–36^ First, in a *derivation cohort* of the initial 1,700 patients enrolled in the trial, we will compare the performance of candidate effect models using 5-fold cross-validation. Specifically, we will consider X-learners with elastic net and random forest base-learners, S-learners with random forest and conditional random forest (cforest) base-learners, T-learner with Bayesian Adaptive Regression Trees (BART) as the base-learner, R-learner with Extreme Gradient Boosting (Xgboost) as the base-learner, and causal forest. The algorithm and base-learner combination resulting in the highest qini coefficient in the out-of-sample predictions from the 5-fold cross-validation will then be fit using all the derivation data to create a final effect model.

Second, in a *validation cohort* of the final 664 patients enrolled in the trial, we will validate the performance of the individualized treatment effect model. To do this, we will use the model to predict the effect of ketamine vs etomidate on 28-day in-hospital mortality for each of the 664 patients in the validation cohort (the patient’s predicted individualized treatment effect at the time of enrolment). We will compare the treatment effect predicted by the model to the treatment effect observed in the validation cohort using the qini coefficient,^37^ the C statistic for benefit,^38^ and measures of clinical utility, in alignment with the PATH guidelines (examining characteristics by quantile of predicted benefit). The effect-modeling analyses may be presented separately from the other trial results if the time required to conduct them or the space required to present them requires doing so.

### 12. Safety Monitoring and Adverse Events

Ensuring patient safety is an essential component of the RSI trial. Both ketamine and etomidate have been approved by the Food and Drug Administration and used in clinical practice for decades with an established safety profile.^39,40^ However, any trial conducted during a high-risk, time-sensitive procedure like tracheal intubation of critically ill patients raises unique safety considerations. This RSI trial addresses these considerations through:

1. Exclusion criteria designed to prevent enrollment of patients likely to experience adverse events from ketamine or etomidate;
2. Systematic collection of safety outcomes relevant to use of ketamine and etomidate in this setting;
3. Structured reporting of adverse events.

### 13. Plan for Communication of Protocol Changes

Any changes to the trial protocol, including changes to eligibility, outcomes, and analyses, will be implemented via a new version of the full trial protocol, tracked with the date of the update, and the version number of the trial protocol. A list summarizing the changes made with each protocol revision will be included at the end of each protocol. The updated protocol will be sent to the sIRB for approval and tracking prior to implementation of the protocol change. At the time of publication, the original trial protocol and the final trial protocol, including the summary of changes made with each protocol version, will be included in the supplementary material for publication.

### 14. Patient Privacy, Data Storage, and Sharing

Federal regulations 45 CFR 46 111 (a) (7) requires that, when appropriate, there are adequate provisions to protect the privacy of patients and to maintain the confidentiality of data. At no time during the course of this study, its analysis, or its publication will patient identities be revealed in any manner. The minimum necessary data containing patient or provider identities will be collected. All patients will be assigned a unique study ID number for tracking. All data collected for this study will be entered into a secure online database. Tools within the secure online database will be used so that only the coordinating center and investigators from the enrolling site will have access to data from patients enrolled at that site. All data will be maintained in the secure online database until the time of study publication. At the time of publication, a de-identified version of the database will be generated. Deidentified data will be available for sharing following trial publication with the requirements and terms of data sharing specified in the trial results manuscript.

### 15. Trial Status

The RSI trial is a pragmatic, multi-center, non-blinded randomized clinical trial comparing the use of ketamine vs the use of etomidate for induction of anesthesia during emergency tracheal intubation of critically ill adults. Enrollment began on April 6, 2022, and is expected to conclude in 2025.

### 16. Supplementary Tables

**Supplementary Table 1. Example of the table of baseline characteristics for the results manuscript.**

| **Table 1. Characteristics of the Patients at Baseline** | | |
| --- | --- | --- |
| **Characteristic** | **Ketamine**  **(N=xxx)** | **Etomidate**  **(N=xxx)** |
| Age, years – median (IQR)* |  |  |
| Female sex – no. (%) |  |  |
| Race or ethnic group – no. (%) ** |  |  |
| Non-Hispanic White |  |  |
| Non-Hispanic Black |  |  |
| Hispanic |  |  |
| Other |  |  |
| Not reported |  |  |
| Weight, kg – median (IQR) |  |  |
| Body mass index – median (IQR) *** |  |  |
| Location of intubation – no. (%) |  |  |
| Emergency department |  |  |
| Intensive care unit |  |  |
| Chronic conditions – no. (%) |  |  |
| Adrenal insufficiency or chronic receipt of corticosteroids |  |  |
| Cirrhosis |  |  |
| Congestive heart failure |  |  |
| Coronary artery disease |  |  |
| Hypertension |  |  |
| Malignancy **** |  |  |
| Acute conditions – no. (%) ***** |  |  |
| Acute cardiac condition ****** |  |  |
| Acute respiratory failure |  |  |
| Acute neurologic condition ******* |  |  |
| Sepsis or septic shock |  |  |
| Glasgow Coma Scale score – median (IQR) |  |  |
| APACHE II score – median (IQR) ******** |  |  |
| Measurement or treatment within the hour before enrollment |  |  |
| Highest heart rate, bpm – median (IQR) |  |  |
| Lowest systolic blood pressure, mm Hg – median (IQR) |  |  |
| Vasopressor receipt – no. (%) |  |  |

* IQR denotes interquartile range.

* Race and ethnicity were reported by patients or their surrogates as part of clinical care and collected from the electronic medical record by research personnel using fixed categories.

*** Body-mass index is computed by the weight in kilograms divided by the square of the height in meters.

**** Malignancy is defined as pulmonary/pleural malignancy, active liquid (leukemia or lymphoma), or malignancy solid non-pulmonary.

***** Data on acute conditions were abstracted from the electronic health record and grouped into prespecified categories. Patients could have had more than one active condition.

****** Acute cardiac condition is defined as cardiac arrest, cardiogenic shock, congestive heart failure, cardiogenic pulmonary edema, pulmonary hypertension, or myocardial infarction present at the time of enrolment.

******* Acute neurologic condition is defined as intracranial bleeding, meningitis or encephalitis, or stroke present at the time of enrolment.

******** Scores on the Acute Physiology and Chronic Health Evaluation (APACHE) II range from 0 to 71, with higher scores indicating a greater severity of illness.

**Supplementary Table 2. Example of the table of characteristics of the intubation procedure for the results manuscript.**

| **Table 2. Characteristics of the Intubation Procedure** | | | |
| --- | --- | --- | --- |
| **Characteristic** | **Ketamine (N=xxx)** | **Etomidate (N=xxx)** | **Difference**  **(95% CI)** |
| Sedative – no. (%) |  |  |  |
| Ketamine |  |  |  |
| Etomidate |  |  |  |
| Other |  |  |  |
| None |  |  |  |
| Neuromuscular blocking agent – no. (%) |  |  |  |
| Rocuronium |  |  |  |
| Succinylcholine |  |  |  |
| None |  |  |  |
| Measurements or treatments at induction of anesthesia |  |  |  |
| Oxygen saturation – median (IQR) |  |  |  |
| Preoxygenation – no. (%) |  |  |  |
| Systolic blood pressure, mm Hg – median (IQR) |  |  |  |
| Vasopressor bolus or increase – no. (%) |  |  |  |
| Laryngoscope – no. (%) |  |  |  |
| Video |  |  |  |
| Direct |  |  |  |
| Instrument used on the first attempt — no. (%) |  |  |  |
| Endotracheal tube with stylet |  |  |  |
| Bougie |  |  |  |

**Supplementary Table 3. Example of the table of outcomes for the results manuscript.**

| **Outcome** | **Ketamine (N=xxx)** | **Etomidate (N=xxx)** | **Difference**  **(95% CI)** |
| --- | --- | --- | --- |
| **Primary outcome** |  |  |  |
| All-cause, 28-day in-hospital mortality – no./total no. (%) |  |  |  |
| **Secondary outcome** |  |  |  |
| Cardiovascular collapse between induction and 2 minutes after intubation – no./total no. (%) |  |  |  |
| Systolic blood pressure < 65 mmHg |  |  |  |
| New or increased vasopressor receipt |  |  |  |
| Cardiac arrest not resulting in death within 1 hour of induction |  |  |  |
| Cardiac arrest resulting in death within 1 hour of induction |  |  |  |
| **Procedural outcomes** |  |  |  |
| Successful intubation on the first attempt – no./total no. (%) |  |  |  |
| Median time from induction to intubation (IQR) – seconds |  |  |  |
| Lowest oxygen saturation, % – median (IQR) |  |  |  |
| Lowest oxygen saturation <80% – no./total no. (%) |  |  |  |
| Systolic blood pressure > 180 mm Hg – no./total no. (%) |  |  |  |
| **Safety outcomes** |  |  |  |
| Systolic blood pressure at 24 hours, mm Hg – median (IQR) |  |  |  |
| Receipt of vasopressors at 24 hours – no./total no. (%) |  |  |  |
| Cardiac arrest between induction of anesthesia and hospital discharge – no./total no. (%) |  |  |  |
| **Clinical outcomes*** |  |  |  |
| Ventilator-free days – median (IQR) |  |  |  |
| Vasopressor-free days – median (IQR) |  |  |  |
| ICU-free days – median – median (IQR) |  |  |  |

### 17. Appendices

#### B. Example Informed Consent Document.

An example of the informed consent document used by trial personnel to obtain prospective, written informed consent for participation in the RSI trial in instances when a patient or Legally Authorized Representative is able to provide informed consent is shown below.

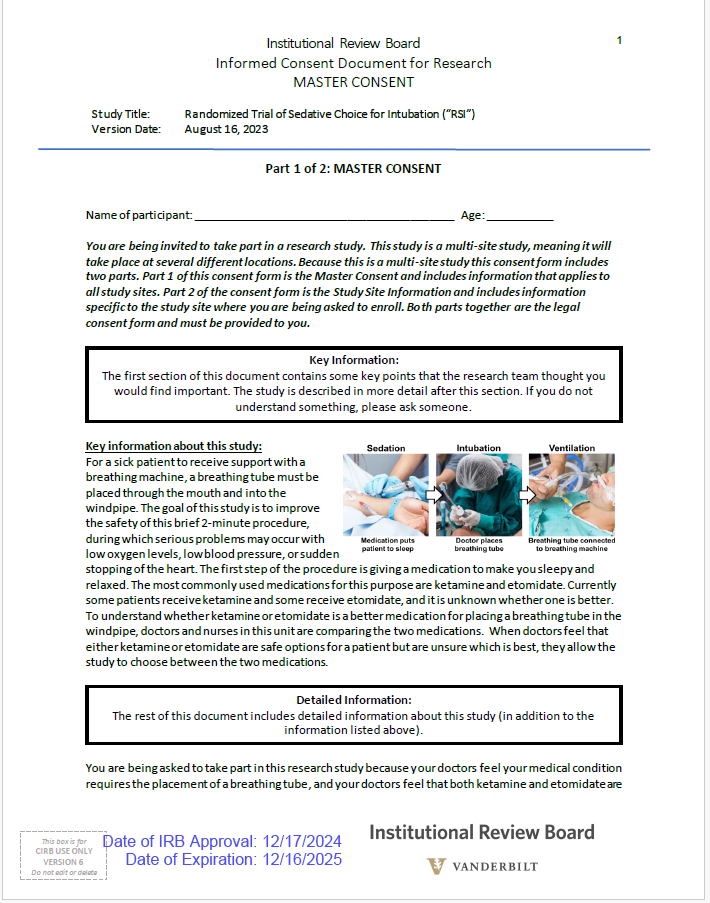

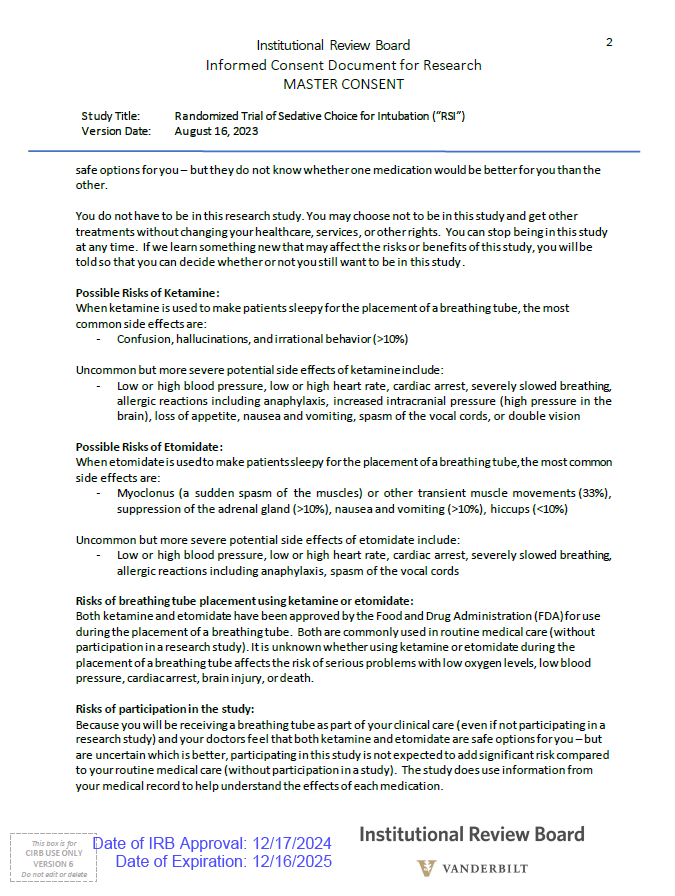

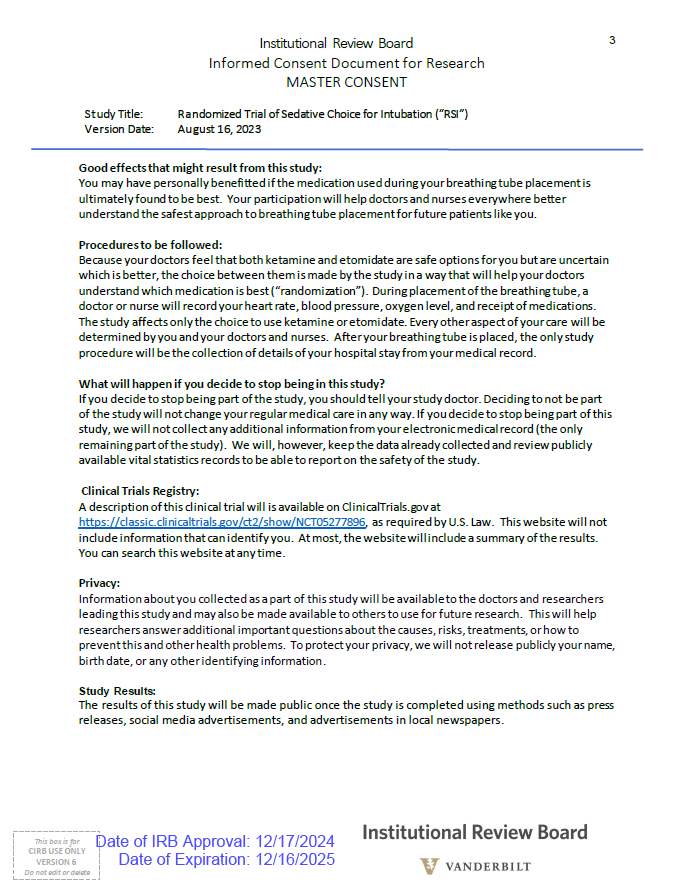

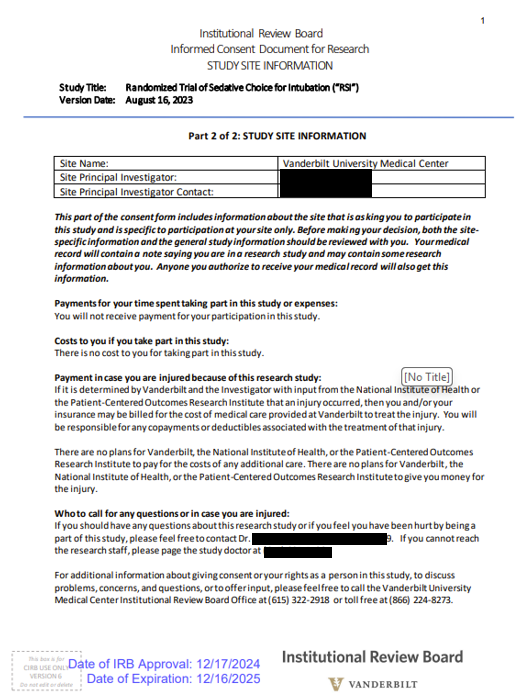

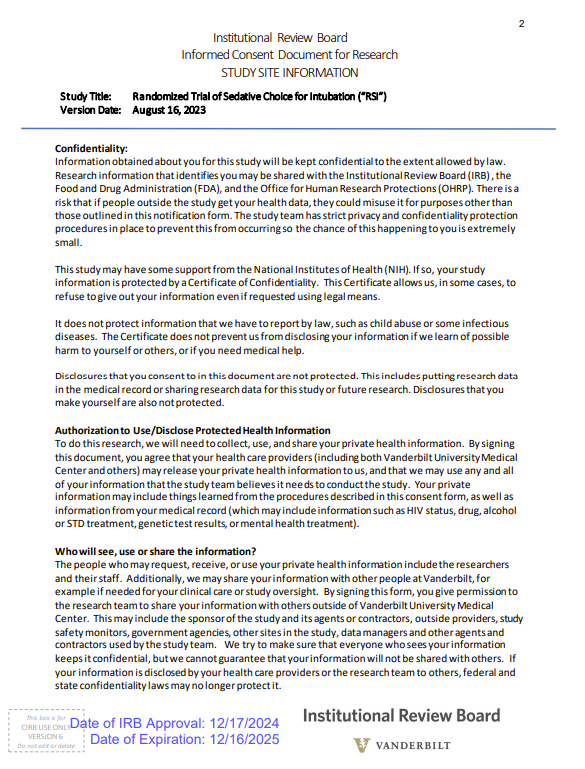

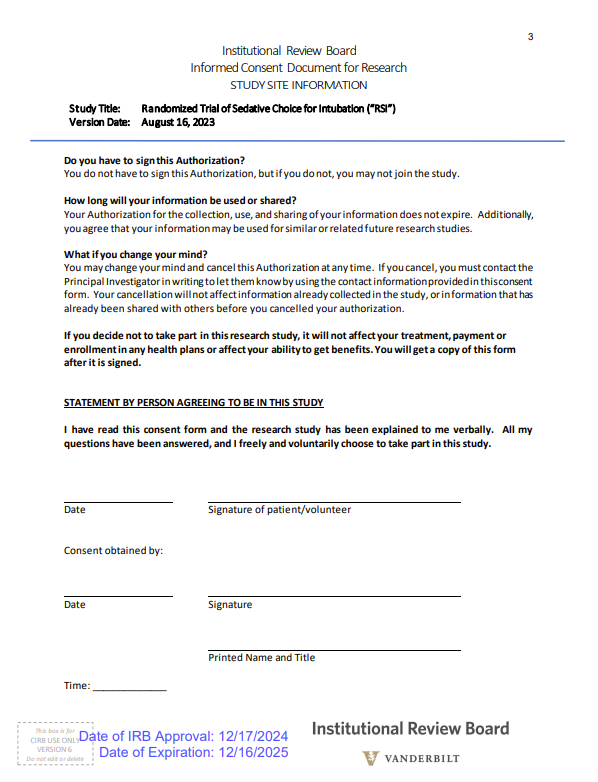

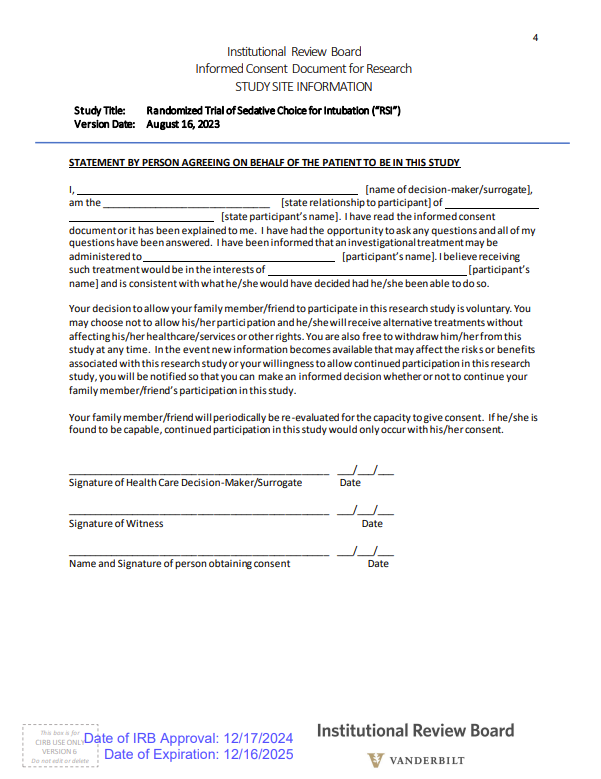

#### B. Example Notification Document.

For patients for whom neither the patient nor a Legally Authorized Representative were able to provide prospective, written informed consent for participation in the RSI trial prior to enrolment, an example of the notification document used to inform the participant or a Legally Authorized Representative of family member of the patient’s enrolment in the trial is shown below.

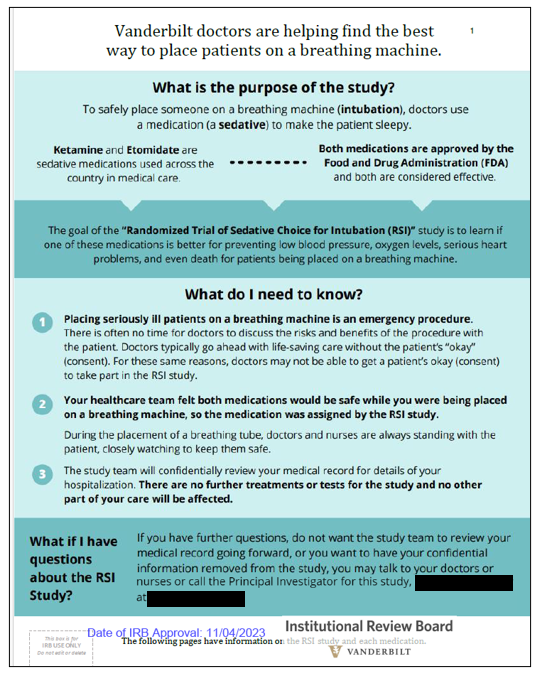

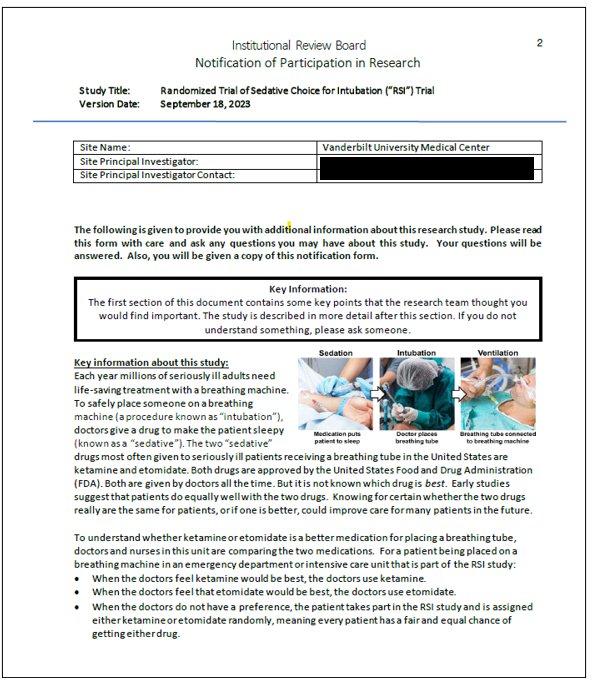

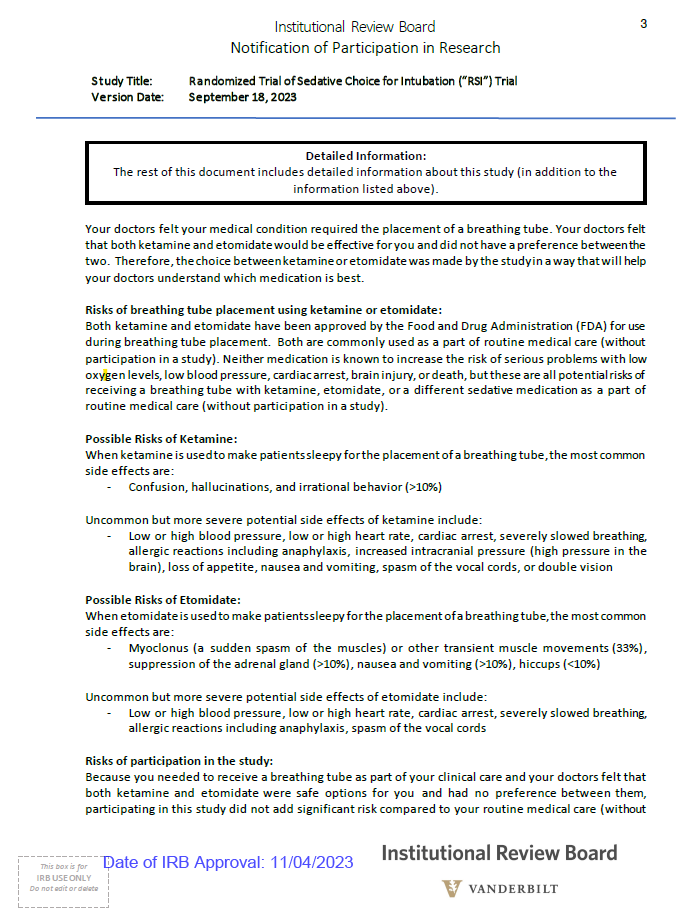

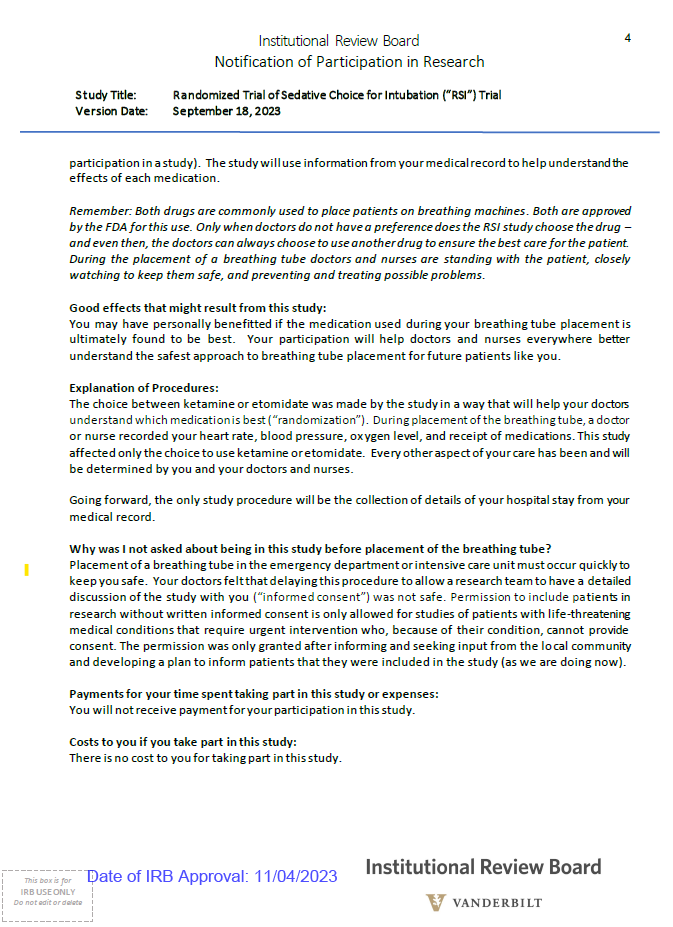

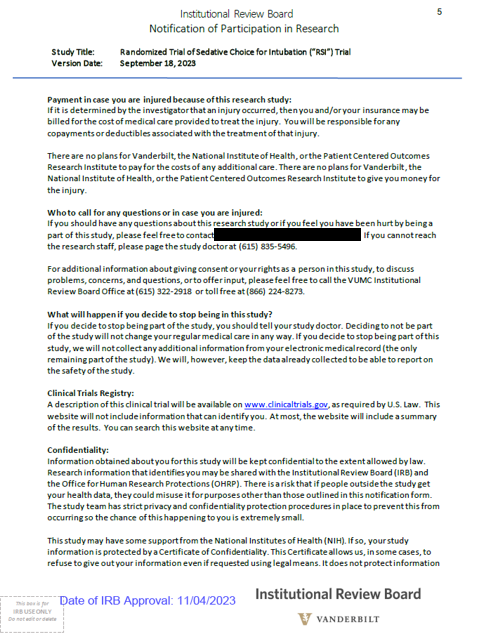

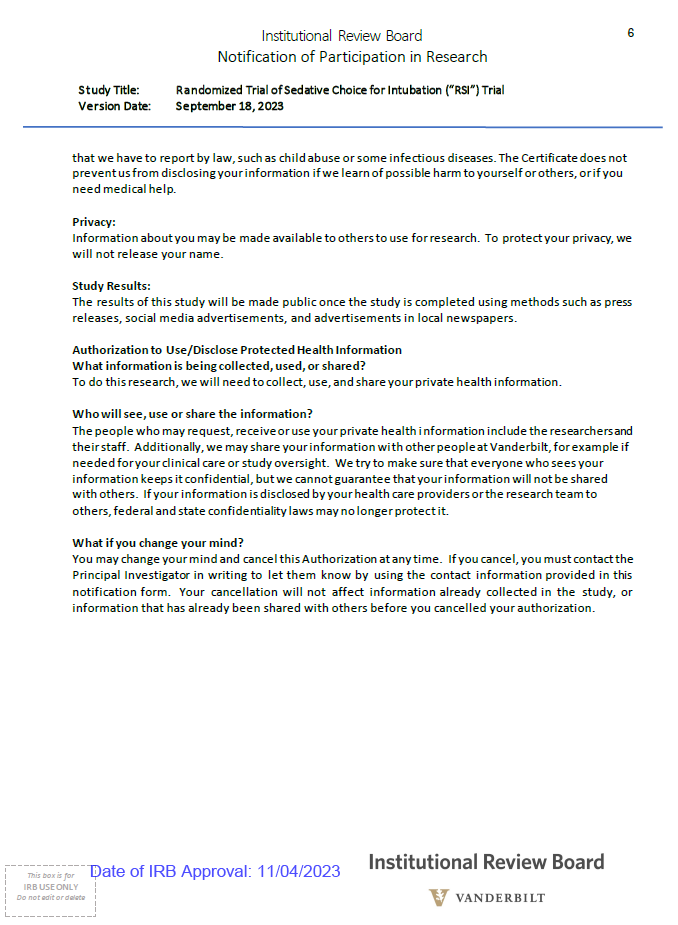

### 18. References

1.	FDA Exception from Informed Consent Requirements for Emergency Research (EFIC) Guidance for Institutional Review Boards, Clinical Investigators, and Sponsors. Date accessed: 01/03/2025. https://www.fda.gov/regulatory-information/search-fda-guidance-documents/exception-informed-consent-requirements-emergency-research

2.	Gibbs KW, Semler MW, Driver BE, et al. Noninvasive Ventilation for Preoxygenation during Emergency Intubation. *N Engl J Med*. 2024;390(23):2165-2177. doi:10.1056/NEJMoa2313680

3.	Prekker ME, Driver BE, Trent SA, et al. Video versus Direct Laryngoscopy for Tracheal Intubation of Critically Ill Adults. *N Engl J Med*. 2023;389(5):418-429. doi:10.1056/NEJMoa2301601

4.	Yarnell CJ, Abrams D, Baldwin MR, et al. Clinical trials in critical care: can a Bayesian approach enhance clinical and scientific decision making? *Lancet Respir Med*. Published online November 20, 2020. doi:10.1016/S2213-2600(20)30471-9

5.	Jabre P, Combes X, Lapostolle F, et al. Etomidate versus ketamine for rapid sequence intubation in acutely ill patients: a multicentre randomised controlled trial. *Lancet Lond Engl*. 2009;374(9686):293-300. doi:10.1016/S0140-6736(09)60949-1

6.	Dinglas VD, Chessare CM, Davis WE, et al. Perspectives of survivors, families and researchers on key outcomes for research in acute respiratory failure. *Thorax*. 2018;73(1):7-12. doi:10.1136/thoraxjnl-2017-210234

7.	Turnbull AE, Sepulveda KA, Dinglas VD, Chessare CM, Bingham CO, Needham DM. Core Domains for Clinical Research in Acute Respiratory Failure Survivors: An International Modified Delphi Consensus Study. *Crit Care Med*. 2017;45(6):1001-1010. doi:10.1097/CCM.0000000000002435

8.	Needham DM, Sepulveda KA, Dinglas VD, et al. Core Outcome Measures for Clinical Research in Acute Respiratory Failure Survivors. An International Modified Delphi Consensus Study. *Am J Respir Crit Care Med*. 2017;196(9):1122-1130. doi:10.1164/rccm.201702-0372OC

9.	Dinglas VD, Faraone LN, Needham DM. Understanding patient-important outcomes after critical illness: a synthesis of recent qualitative, empirical, and consensus-related studies. *Curr Opin Crit Care*. 2018;24(5):401-409. doi:10.1097/MCC.0000000000000533

10.	Hodgson CL, Turnbull AE, Iwashyna TJ, et al. Core Domains in Evaluating Patient Outcomes After Acute Respiratory Failure: International Multidisciplinary Clinician Consultation. *Phys Ther*. 2017;97(2):168-174. doi:10.2522/ptj.20160196

11.	Mikkelsen ME, Still M, Anderson BJ, et al. Society of Critical Care Medicine’s International Consensus Conference on Prediction and Identification of Long-Term Impairments After Critical Illness. *Crit Care Med*. 2020;48(11):1670-1679. doi:10.1097/CCM.0000000000004586

12.	Spruit MA, Holland AE, Singh SJ, Tonia T, Wilson KC, Troosters T. COVID-19: Interim Guidance on Rehabilitation in the Hospital and Post-Hospital Phase from a European Respiratory Society and American Thoracic Society-coordinated International Task Force. *Eur Respir J*. Published online August 13, 2020:2002197. doi:10.1183/13993003.02197-2020

13.	Rubin EB, Buehler A, Halpern SD. Seriously Ill Patients’ Willingness to Trade Survival Time to Avoid High Treatment Intensity at the End of Life. *JAMA Intern Med*. 2020;180(6):907-909. doi:10.1001/jamainternmed.2020.0681

14.	Rubin EB, Buehler AE, Halpern SD. States Worse Than Death Among Hospitalized Patients With Serious Illnesses. *JAMA Intern Med*. 2016;176(10):1557-1559. doi:10.1001/jamainternmed.2016.4362

15.	Auriemma CL, Harhay MO, Haines KJ, Barg FK, Halpern SD, Lyon SM. What Matters to Patients and Their Families During and After Critical Illness: A Qualitative Study. *Am J Crit Care Off Publ Am Assoc Crit-Care Nurses*. 2021;30(1):11-20. doi:10.4037/ajcc2021398

16.	Halpern SD, Temel JS, Courtright KR. Dealing With Death as an Outcome in Supportive Care Clinical Trials. *JAMA Intern Med*. 2021;181(7):895-896. doi:10.1001/jamainternmed.2021.1816

17.	Schandelmaier S, Briel M, Varadhan R, et al. Development of the Instrument to assess the Credibility of Effect Modification Analyses (ICEMAN) in randomized controlled trials and meta-analyses. *CMAJ Can Med Assoc J J Assoc Medicale Can*. 2020;192(32):E901-E906. doi:10.1503/cmaj.200077

18.	Singer M, Deutschman CS, Seymour CW, et al. The Third International Consensus Definitions for Sepsis and Septic Shock (Sepsis-3). *JAMA*. 2016;315(8):801-810. doi:10.1001/jama.2016.0287

19.	Cuthbertson BH, Sprung CL, Annane D, et al. The effects of etomidate on adrenal responsiveness and mortality in patients with septic shock. *Intensive Care Med*. 2009;35(11):1868-1876. doi:10.1007/s00134-009-1603-4

20.	Lipiner-Friedman D, Sprung CL, Laterre PF, et al. Adrenal function in sepsis: The retrospective Corticus cohort study: *Crit Care Med*. 2007;35(4):1012-1018. doi:10.1097/01.CCM.0000259465.92018.6E

21.	Tsai MH, Peng YS, Chen YC, et al. Adrenal insufficiency in patients with cirrhosis, severe sepsis and septic shock. *Hepatol Baltim Md*. 2006;43(4):673-681. doi:10.1002/hep.21101

22.	Jabre P, Combes X, Lapostolle F, et al. Etomidate versus ketamine for rapid sequence intubation in acutely ill patients: a multicentre randomised controlled trial. *Lancet Lond Engl*. 2009;374(9686):293-300. doi:10.1016/S0140-6736(09)60949-1

23.	Mohammad Z, Afessa B, Finkielman JD. The incidence of relative adrenal insufficiency in patients with septic shock after the administration of etomidate. *Crit Care Lond Engl*. 2006;10(4):R105. doi:10.1186/cc4979

24.	Albert SG, Ariyan S, Rather A. The effect of etomidate on adrenal function in critical illness: a systematic review. *Intensive Care Med*. 2011;37(6):901-910. doi:10.1007/s00134-011-2160-1

25.	Abani O, Abbas A, Abbas F, et al. Tocilizumab in patients admitted to hospital with COVID-19 (RECOVERY): a randomised, controlled, open-label, platform trial. *The Lancet*. 2021;397(10285):1637-1645. doi:10.1016/S0140-6736(21)00676-0

26.	Srivilaithon W, Bumrungphanithaworn A, Daorattanachai K, et al. Clinical outcomes after a single induction dose of etomidate versus ketamine for emergency department sepsis intubation: a randomized controlled trial. *Sci Rep*. 2023;13(1):6362. doi:10.1038/s41598-023-33679-x

27.	de Jong FH, Mallios C, Jansen C, Scheck PA, Lamberts SW. Etomidate suppresses adrenocortical function by inhibition of 11 beta-hydroxylation. *J Clin Endocrinol Metab*. 1984;59(6):1143-1147. doi:10.1210/jcem-59-6-1143

28.	Wunsch H, Bosch NA, Law AC, et al. Evaluation of Etomidate Use and Association with Mortality Compared with Ketamine Among Critically Ill Patients. *Am J Respir Crit Care Med*. Published online August 22, 2024. doi:10.1164/rccm.202404-0813OC

29.	Kolenda H, Gremmelt A, Rading S, Braun U, Markakis E. Ketamine for analgosedative therapy in intensive care treatment of head-injured patients. *Acta Neurochir (Wien)*. 1996;138(10):1193-1199. doi:10.1007/BF01809750

30.	Sprung J, Schuetz SM, Stewart RW, Moravec CS. Effects of ketamine on the contractility of failing and nonfailing human heart muscles in vitro. *Anesthesiology*. 1998;88(5):1202-1210. doi:10.1097/00000542-199805000-00010

31.	Kongsayreepong S, Cook DJ, Housmans PR. Mechanism of the direct, negative inotropic effect of ketamine in isolated ferret and frog ventricular myocardium. *Anesthesiology*. 1993;79(2):313-322. doi:10.1097/00000542-199308000-00017

32.	Iwashyna TJ, Burke JF, Sussman JB, Prescott HC, Hayward RA, Angus DC. Implications of Heterogeneity of Treatment Effect for Reporting and Analysis of Randomized Trials in Critical Care. *Am J Respir Crit Care Med*. 2015;192(9):1045-1051. doi:10.1164/rccm.201411-2125CP

33.	Knaus WA, Draper EA, Wagner DP, Zimmerman JE. APACHE II: a severity of disease classification system. *Crit Care Med*. 1985;13(10):818-829.

34.	Kent DM, van Klaveren D, Paulus JK, et al. The Predictive Approaches to Treatment effect Heterogeneity (PATH) Statement: Explanation and Elaboration. *Ann Intern Med*. 2020;172(1):W1-W25. doi:10.7326/M18-3668

35.	Seitz KP, Spicer AB, Casey JD, et al. Individualized Treatment Effects of Bougie vs Stylet for Tracheal Intubation in Critical Illness. *Am J Respir Crit Care Med*. Published online March 6, 2023. doi:10.1164/rccm.202209-1799OC

36.	Buell KG, Spicer AB, Casey JD, et al. Individualized Treatment Effects of Oxygen Targets in Mechanically Ventilated Critically Ill Adults. *JAMA*. 2024;331(14):1195-1204. doi:10.1001/jama.2024.2933

37.	Belbahri M, Murua A, Gandouet O, Nia VP. Uplift Regression: The R Package tools4uplift. :22.

38.	van Klaveren D, Steyerberg EW, Serruys PW, Kent DM. The proposed “concordance-statistic for benefit” provided a useful metric when modeling heterogeneous treatment effects. *J Clin Epidemiol*. 2018;94:59-68. doi:10.1016/j.jclinepi.2017.10.021

39.	FDA Ketamine. Date Accessed: 11/12/2024. https://www.accessdata.fda.gov/drugsatfda_docs/label/2018/016812s040lbl.pdf

40.	FDA Etomidate. Date Accessed: 11/12/2024. November 12, 2024. https://www.accessdata.fda.gov/drugsatfda_docs/label/2017/018227s032lbl.pdf
